# CanVar-UK: an online data platform supporting collaborative diagnostic interpretation of germline variants in cancer susceptibility genes

**DOI:** 10.64898/2026.03.23.26349065

**Authors:** Charlie F. Rowlands, Subin Choi, Sophie Allen, Zeid Kuzbari, Cankut Çubuk, Razvan Sultana, Beth Torr, Miranda Durkie, George J. Burghel, Rachel Robinson, Alison Callaway, Joanne Field, Bethan Frugtniet, Sheila Palmer-Smith, Jonathan Grant, Judith Pagan, Elizabeth Johnston, Trudi McDevitt, Lowri Hughes, Laura Yarram-Smith, Peter Logan, Laura Reed, Katie Snape, Terri McVeigh, Helen Hanson, Alice Garrett, Clare Turnbull, CanVIG-UK

**Affiliations:** Division of Genetics and Epidemiology, The Institute of Cancer Research, London, UK; Centre for Experimental Medicine & Rheumatology, William Harvey Research Institute, Barts and The London School of Medicine and Dentistry, Queen Mary University of London, London, UK; Barts Health NHS Trust and National Institute for Health and Care Research (NIHR) Barts Biomedical Research Centre (BRC), London, UK; Garvan Institute of Medical Research, Sydney, Australia; North East and Yorkshire Genomic Laboratory Hub, Sheffield Children’s NHS Foundation Trust, Sheffield, UK; Division of Cancer Sciences, School of Medical Sciences, Faculty of Biology, Medicine and Health, The University of Manchester, Manchester, UK; Manchester Centre for Genomic Medicine and NW Laboratory Genetics Hub, Manchester University Hospitals NHS Foundation Trust, Manchester, UK; The Leeds Genetics Laboratory, NEY Genomic Laboratory Hub, Leeds Teaching Hospitals NHS Trust, Leeds, UK; Central and South Genomics Laboratory Hub, Wessex Genomics Laboratory Service, University Hospital Southampton NHS Foundation Trust, Salisbury, UK; Genomics and Molecular Medicine Service, Nottingham University Hospitals NHS Trust, Nottingham, UK; St George’s University Hospitals NHS Foundation Trust, Tooting, London, UK; Wales Genomic Health Centre, Cardiff and Vale University Health Board, Cardiff, UK; West of Scotland Centre for Genomic Medicine, Queen Elizabeth University Hospital, NHS Greater Glasgow and Clyde, Glasgow, UK; South East Scotland Clinical Genetics, Western General Hospital, Edinburgh, UK; Department of Clinical Genetics, CHI at Crumlin, Dublin, Ireland; West Midlands Genomics Laboratory, Birmingham Women’s and Children’s NHS Foundation Trust, Birmingham, UK; North Bristol NHS Trust, Southmead Hospital, Bristol, UK; Belfast Health and Social Care Trust, Royal Victoria Hospital, Belfast, UK; Rare & Inherited Disease Laboratory, NHS North Thames Genomic Laboratory Hub, Great Ormond Street Hospital for Children NHS Foundation Trust, London, UK; The Royal Marsden NHS Foundation Trust, Fulham Road, London; Peninsula Regional Genetics Service, Royal Devon University Healthcare NHS Foundation Trust, Exeter, UK; Department of Clinical and Biomedical Sciences, University of Exeter Medical School, Exeter, United Kingdom

## Abstract

Interpretation of germline variants in cancer susceptibility genes (CSGs) requires the collation of variant-level data from diverse sources, as well as the assembly of comprehensive clinical data, often necessitating sharing of information between genomic testing centers. Although a number of variant interpretation tools exist, there remains a need for a CSG-focused platform tailored to the diverse range of ClinGen variant curation expert panel guidance in these difficult-to-interpret genes. Here, we describe CanVar-UK, a freely-accessible web platform to assist in the interpretation of germline CSG variants. CanVar-UK contains variant-level data for over 1.7 million single nucleotide variants, comprising all coding variants in 115 established CSGs. These data include: *in silico* scores from 11 tools of clinical relevance; population allele frequencies from gnomAD v4.1 and case counts from NHS genomic testing via linkage to the National Disease Registration Service; variant-level readouts from 31 different functional and splicing studies across 13 CSGs; genetic epidemiology studies of the *BRCA1/2* genes; and live linkage to existing consensus classifications in the ClinVar database. CanVar-UK additionally has a diagnostic discussion forum functionality, via which users are able to email the rest of the user base with queries and/or suggested classifications, facilitating the exchange of clinical and classification data between diagnostic centers. Already widely used by the NHS clinical workforce in the CSG space (with 879 registered NHS users), CanVar-UK has a rapidly growing international user base, with 607 registered users based outside the UK. We believe CanVar-UK to be an invaluable resource for germline CSG variant interpretation.

## Main text

Cancer susceptibility genetics is a sizeable subspecialty of clinical genetics, typically constituting a quarter to a half of overall genetic testing activity^1^. In the UK, there are >100 cancer susceptibility genes included on the NHS National Test Directory^2^. Diagnostic testing in cancer patients is expanding from only patients judged as having a high likelihood of pathogenic variant identification based on their personal and family history of cancer to a mainstream offering in a wider group of patients. Many germline pathogenic variants are identified when running large tumor panels to inform oncological management^3^. Alongside return of incidental/secondary findings and direct-to-consumer testing, there is a wider transition from diagnostic testing of cancer susceptibility genes (CSGs) in the medical setting to population-level screening^4^. A recent study estimated that 5.05% of the population carry a pathogenic variant in a cancer susceptibility gene (20,968/414,830 individuals in the US-based All of Us dataset)^5^.

Accurate variant interpretation and classification are fundamental to any application of laboratory genomic analyses for clinical care. Inaccurate classifications - either erroneously classifying a variant as (likely) pathogenic or failing to classify a (likely) pathogenic variant as such - can result in clinical harms. Unlike in many rare diseases, most germline pathogenic variants in CSGs are autosomal dominant and typically inherited, rather than arising *de novo*. Accordingly, wide cascade testing of families is typical, thus necessitating sharing of variant details and classifications nationally and internationally. Clinical actions, such as risk-reducing surgery, taken on the basis of CSG variant classifications may be irreversible. For all these reasons, inconsistent variant classification and downstream reclassification of CSG variants can be highly problematic.

Released in 2015, version 3.0 of the ACMG/AMP variant classification framework represented an important shift towards a single internationally standard approach to variant classification^6^. Following its endorsement in 2016 by the UK Association of Clinical Genomic Science (UK-ACGS), Cancer Variant Interpretation Group UK (CanVIG-UK) was initiated in 2017 as a subspecialty multidisciplinary network of NHS clinical scientists and genetics clinicians to provide training, education and embedding of the new ACMG/AMP framework, with the goal of improving national consistency in classification of variants in CSGs^7-9^. CanVIG-UK has continued to convene monthly, (i) evolving detailed consensus and gene-specific variant classification guidance, (ii) generating new methods for quantitation of evidence of pathogenicity/benignity, (iii) developing themed consensus national approaches for emerging issues such as reclassification and reduced penetrance variants, and (iv) providing a nexus for interaction and information flow between the UK cancer susceptibility diagnostic community and international groups, including ClinGen and the AVE Alliance^8,10-20^.

Classification of genomic variants requires assembly of a wealth of different evidence types from potentially disparate sources, which include predictions from *in silico* tools, variant frequencies from population databases, results from functional assays and clinical observations. Some of these resources may be readily publicly accessible, some may require logging into specific websites, and some may be buried in supplementary tables from manuscripts. ClinVar provides an invaluable international repository of variant classifications. Nevertheless, while individual submissions may delineate applied evidence, ClinVar itself does not centralize the constituent data resources^21^ and it is the recommendation of ClinGen that, even for 3-star (Variant Curation Expert Panel, or VCEP) classifications, the variant should be reviewed and re-classified upon each clinical observation, in case of emergence of new data and/or any errors upstream of ClinVar deposition^22,23^. Assembly of sufficient clinical observations aligned to phenotypic data is typically the major challenge for classification of most variants: this has to date often necessitated lengthy iterative email communication across different laboratory professionals and clinicians in order to quantify and assess phenotypic findings linked to previous instances of the variant.

In parallel with the inception of the group in 2017, we sought to develop a digital platform to support the CanVIG-UK community bespoke for CSGs, with the aim of improving quality, consistency and efficiency of variant classification. The platform, named CanVar-UK (https://canvaruk.org/), is built using the PHP-based Laravel framework (Supplemental Methods) and contains data on 1,744,860 variants from 116 CSGs, comprising all possible single nucleotide variants in the coding sequence (±25 bp into the introns/untranslated regions) of clinically selected transcripts (Table S1). Indels and CNVs have subsequently been added to the database when clinically observed and submitted by a user, thus allowing posting of queries and classifications for the variant via the diagnostic discussion forum (see below). A dedicated email inbox allows rapid response to CanVar-UK user queries, facilitating the design and implementation of new features and the addition of new *in silico*, functional and genetic epidemiology datasets as per user-community requests, as well as agile bug fixes.

In our iterative development of the CanVar-UK platform since 2017, we have sought to address the following objectives:

### (i) To provide variant-level data in an intuitive, ordered, rapidly-accessible format

We designed an interface for presentation of variant-level data with six tabs: (i) summary, *in silico* predictions, (iii) case/control frequency, (iv) genetic epidemiology (v) functional/splicing assays and (vi) the CanVar-UK discussion forum (Figure 1). The user selects the variant under evaluation using a drop-down of genes (with commonly interrogated genes at the top), and then enters the variant name (via p. or c. nomenclature with predictive matching provided). Tabs (i)-(v) can be accessed directly without any log-in.

**Figure 1.**
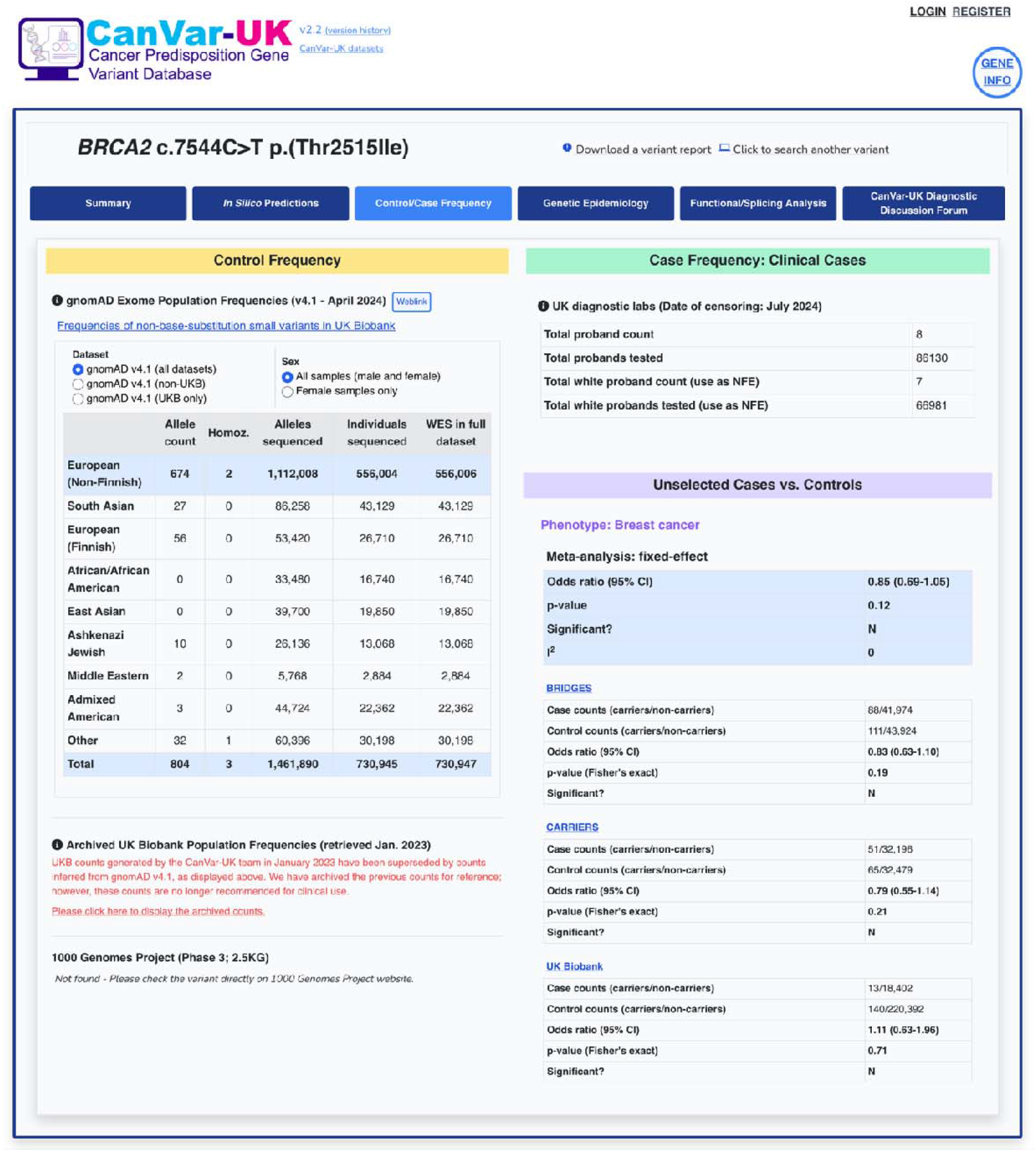
CanVar-UK case and control frequency interface for exemplar variant BRCA2 c.7544C>T. Shown is an illustrative example of a CanVar interface for case/control frequency. CanVar-UK displays allele counts from the gnomAD v4.1 population dataset and allows stratification of counts according to both the constituent gnomAD datasets (UK Biobank (UKB) vs. non-UKB) and sex of samples (all samples or female-only). Variant counts from Phase 3 of the 1000 Genomes Project are also displayed. Collated counts of variant observations and total proband tests from NHS diagnostic testing centers are shown, allowing quantification of case-control signal. For selected hereditary breast and ovarian cancer susceptibility genes, CanVar-UK additionally displays estimated variant effect size from three unselected case-control datasets: the BRIDGES^25^ and CARRIERS^26^ studies, plus UK Biobank. Where the variant is identified in multiple of these datasets, a pre-computed meta-analyzed predicted effect size is also shown.

### (ii) To provide wide-ranging resources of variant-level data that are generic across cancer susceptibility genes (“pan-gene” resources)

Variant-level annotations presented include multiple *in silico* prediction tools, specifically those widely used and/or recommended by VCEPs. Information bubbles are provided advising on their provenance, application and thresholds (Figures S1-2). Live ClinVar classifications are dynamically retrieved for each variant via the ClinVar API. A “PS1/PM5 lookup” function also allows retrieval of ClinVar classifications for the other eight base substitutions of the codon (along with their REVEL scores; Figure S1). Interrogation of Google and Google Scholar is facilitated via automated generation of the relevant Boolean search term (e.g.”*BRCA1*”(“c.5207T>C”|”c.5207T/C”|”CM041712”|”Val1736Ala”|”V1736A”)). Links to LOVD, cBioPortal and VarSome are also provided.

### (iii) To curate and present tailored resources of variant-level data relevant to specific genes (gene-specific resources)

Variant-level outputs from functional assays, both low-throughput and multiplexed assays of variant effect (MAVEs), are presented, in particular those recommended by VCEPs (Figure S3; Table S2). Explanatory information bubbles are provided delineating assay methodology and scoring thresholds. Data from analyses of splicing function are also included: this includes both published series of splicing analyses and data shared by the UK laboratories undertaking cDNA splicing analyses. Key multifactorial genetic epidemiology analyses are included for *BRCA1* and *BRCA2*, with summary and constituent (log) likelihood ratios presented, along with explanatory information bubbles (Figure S4; Table S3).

### (iv) To house and present summary variant counts from UK NHS laboratory and other case cohort testing alongside population frequency estimates

Since 2017, linkage with historic and prospective submission to the NHS National Disease Registration Service (NDRS) and CanVar-UK has been established, comprising patient-level counts of variant observations from all 17 diagnostic laboratories in England analyzing CSGs^24^. The summary counts are released bi-annually by NDRS for presentation on CanVar-UK, stratified by high-level ancestry, with the corresponding denominator of tests performed (Figure 1). Registered users with an NHS email address are further able to view variant counts stratified by the specific testing laboratory.

Variant frequency from population-level datasets such as gnomAD, UK Biobank and 1000 Genomes are presented, stratified by ancestry group. We provide facility to toggle between all participants and female-only participants (as per desired control group). Together, these resources inform calculation of case-control evidence weighting (PS4; Figure 1).

With permission from collaborators, we have also recently curated variant-level frequencies from two large-scale sequencing projects (BRIDGES^25^ and CARRIERS^26^) comprising unselected breast cancer cases and control series, presenting these in combination with breast cancer cases and controls from UK Biobank (Figure 1).

### (v) To present relevant gene-level resources to complement variant-level resources

We present key gene-level data informative to variant interpretation, such as the full set of exon and intron sizes and boundaries and information on transcripts. We also present relevant gene-specific variant classification guidance, with relevant links to and details of both VCEP and CanVIG-UK resources (Figure S5).

### (vi) To create a dynamic user forum for communication and information-sharing amongst diagnostic users

Following registration and login, a user is able to share classifications, information and queries relating to the variant under interrogation via the “CanVar-UK Diagnostic Discussion Forum”. The user can select whether their communication against the variant is captured just as a stored post or whether they wish to also issue an email to the diagnostic discussion forum membership (Figure S6). The email is structured with the variant name in the header and labelled as being either “standard” or “urgent”. A recipient can respond via the email’s embedded reply function and their responses are likewise captured in the forum log. Users can also upload documents against a variant to the CanVar-UK Diagnostic Discussion Forum, thus enabling ordered storage and community-wide access to data, additional emails, and slides which have underpinned discussions and classifications. It is explicitly recommended that any shared clinical information is de-identified. We have added a facility to flag whether a classification of reduced penetrance was considered and/or reached during variant interpretation^15^.

### (vii) A variant-level report output

Users can download a PDF report for the variant under interrogation. This report contains all the annotations for the variant from general, *in silico*, case-control, functional/splicing and genetic epidemiology resources (Figure S7).

CanVar-UK is publicly accessible and can be interrogated without login access. Only the CanVar-UK diagnostic discussion forum area and stratified UK laboratory counts are password-controlled. All features, with and without login requirements, are free to access. Previously access was restricted to NHS clinical users, but since 2022, users from an established diagnostic laboratory can apply for access to the CanVar-UK diagnostic user forum; access is granted pending manual confirmation of their clinical diagnostic credentials. As of the end of 2025, CanVar-UK has 1536 individuals registered for access to the diagnostic discussion forum: 879 NHS members, 50 non-NHS UK members and 607 international members. Over 1292 users are from Europe (of whom 929 are UK-based), 79 from South America, 63 from North America, 57 from Oceania, 39 from Asia and 6 from Africa (Figures 2a-b). In 2025, over 500 email enquiries were sent out via the diagnostic user forum, with an average of approximately 42 per month (Figure 2c). The most common genes for which forum posts are submitted are those associated with hereditary breast and ovarian cancer (HBOC; BRCA1, BRCA2, PALB2, ATM, CHEK2, RAD51C, RAD51D and BRIP1) and mismatch repair (MLH1, MSH2, MSH6 and PMS2).

**Figure 2.**
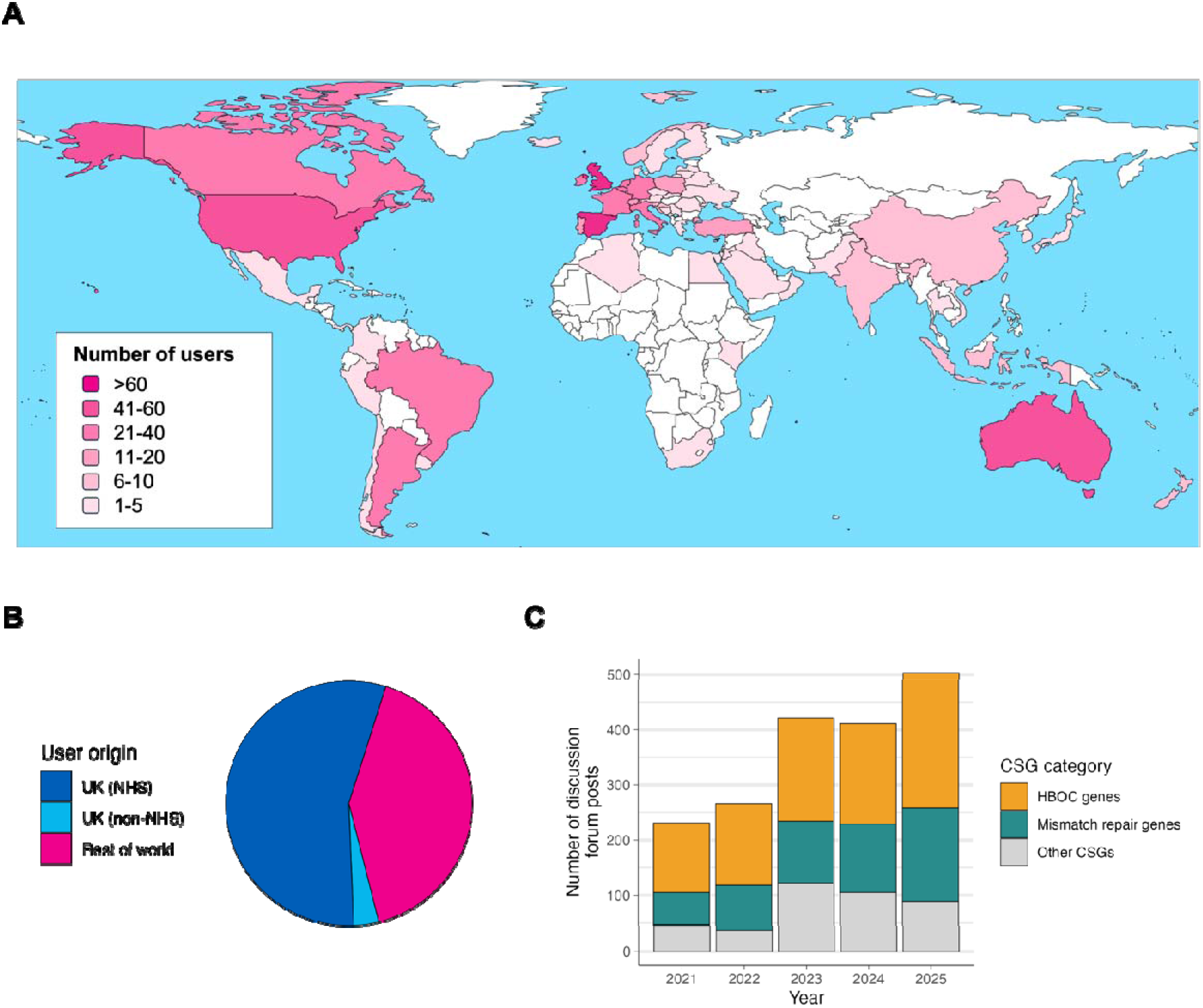
Membership and diagnostic forum usage statistics for CanVar-UK. As of 12/31/2025, (**A**) CanVar-UK membership comprises 1536 individuals from 57 countries across six continents. (**B**) Breakdown of CanVar-UK users: 57.2% (879/1536) NHS registered staff; 3.3% (50/1536) non-NHS users based in the UK; 38.2% (607/1536) international users. (**C**) Number of posts on the CanVar-UK diagnostic discussion forum against time and breakdown by gene grouping

In a recent survey of CanVIG-UK clinical scientist members (Supplemental Methods; results for all respondents presented in Table S3), respondents were asked to rank the relative utility of CanVar-UK, Alamut (SOPHiA Genetics, Lausanne, Switzerland), ClinVar^27^, Franklin^28^ and VarSome^29^ for assembly of data for variant interpretation: 66.3% (65/98) rated CanVar-UK the most useful, with a further 19.4% ranking it second-most useful of the five tools (Figure S8). 76.5% (75/98) of respondents reported using CanVar-UK at least weekly to access information for variant classification, and 61.2% (60/98) reported posting questions via the discussion forum (Figure S9). When asked to rate the usefulness of 12 individual CanVar-UK features from 0 (“not at all useful”) to 5 (“extremely useful”), 10 features were given a rating of very/extremely useful (4 or 5) by more than half of respondents (Figure S10). When asked the question “Overall, how important is CanVar-UK for your work?”, 86.7% (85/98) participants responded with a score of 8 or higher (0 being “Not at all” and 10 being “Essential”).

We have presented an overview of the design, features and usage of the CanVar-UK platform by the community of NHS clinicians and clinical scientists within the CanVIG-UK community working within cancer susceptibility genomics.

Both in the resources held therein and its operational functionality, CanVar-UK was developed in consultation with the CanVIG-UK clinical user community. Evolving nimbly and responsively to the needs of this community, we have sought to import into CanVar-UK potentially relevant resources as requested by the CanVIG-UK community, comprising both pan-gene and gene-specific resources. A key challenge has thus been prioritizing of developer time in regard to inputting data from the large number of small functional assays and clinical data series, particularly in relation to genes of lesser testing volumes.

Following recent surveying of the CanVIG-UK group, we anticipate that near-future directions for CanVar-UK will include increased incorporation of MAVE data with enhanced concomitant focus on providing additional metadata informing assay-level (and even variant-level) evidence scoring^9^. We shall also continue to use the CanVar-UK platform to support CanVIG-UK guideline development and audit activities, for example capturing of variants for which reduced penetrance is suspected^15^. Additionally, we shall look to align structuring of data and classifications in CanVar-UK with the anticipated ACMG/AMP/CAP/ClinGen v4.0 classification framework.

Although transition to ACMG/ClinGen v4.0 will provide opportunities for refinement and automation of some variant classification codes and management by grouping and capping of potentially orthogonal data, the practice of clinical variant classification will still require complex and often subjective evaluation regarding scoring and combination of different data types^30,31^. The CanVIG-UK national network for cancer susceptibility, supported by the CanVar-UK platform and diagnostic discussion forum, offers a template for how a national subspeciality genomics network might operate to optimize collaborative working in sharing clinical data, evolving practice, educating the community and ratifying individual instances of challenging classifications.

## Data Availability

The CanVar-UK website is freely accessible at https://canvaruk.org/, with full access to website features upon registration and validation of user credentials.

## Author contributions

The CanVar-UK platform was conceived by C.T. and designed, developed and maintained by C.F.R., S.C., C.C. and R.S. Generation and/or processing of data for display on CanVar-UK was conducted by C.F.R., S.C., C.C., R.S., S.A. and B.T. All authors were involved in the suggestion of key features and datasets for the CanVar-UK website. This manuscript was drafted by C.T. and C.F.R., with all authors contributing to review of the manuscript. Funding for the project was obtained by C.T.

## Acknowledgements

Data curation and software engineering for CanVar-UK has been supported by Cancer Research UK Catalyst Award CanGene-CanVar (C61296/A26688, 2019-2024) and by the Cancer Research UK Programme Award CG-MAVE (EDDPGM-Nov22/100004, 2024-2027). A.G. receives funding from NHS England.

