## Supplemental Material for "CanVar-UK: an online data platform supporting collaborative diagnostic interpretation of germline variants in cancer susceptibility genes"

**Supplemental Methods**

*CanVar-UK infrastructure*

The CanVar-UK website is built using the PHP-based Laravel web framework (v8.83.12), with its variant database implemented in MariaDB (v10.4.7). The website is hosted on a virtual server running Ubuntu 20.04, maintained by the Institute of Cancer Research. Bespoke Python scripts are used to generate all possible SNVs for a transcript(s) of interest. *In silico* data are primarily taken from pre-generated genome-wide online scoresets accompanying original publications. Raw data from functional, splicing and genetic epidemiological analyses are processed in-house from original publications to retain columns for display, ensuring transcripts match between the assay and the CanVar-UK. Live ClinVar classifications (including same-codon classifications for the PM5 functionality) are retrieved via the ClinVar API.

*CanVIG-UK survey methodology*

The online survey was developed on the SurveyMonkey platform and was piloted on CStAG members before wider circulation to the CanVIG-UK membership. It comprised 20 questions relating to CanVIG-UK meetings and resources, of which four questions relating specifically to CanVar-UK are presented in this manuscript (Table S3). Survey respondents were CanVIG-UK members who self-selected to participate after a verbal introduction to the survey in the November 2025 CanVIG-UK monthly meeting. A link to the survey was circulated both during the meeting and afterwards by email to all CanVIG-UK members. The survey remained open from 14^th^-28^th^ November 2025.

A total of 137 members completed the survey, of whom 98 were clinical scientists, with their role self-identified as being either “Qualified clinical scientist” or “Clinical scientist trainee”. Figures S8-S11 show only the responses of clinical scientists, with the responses of the full set of respondents detailed in Table S3. Quantitative analyses were performed using R v4.4.1.

**Supplemental Figures**

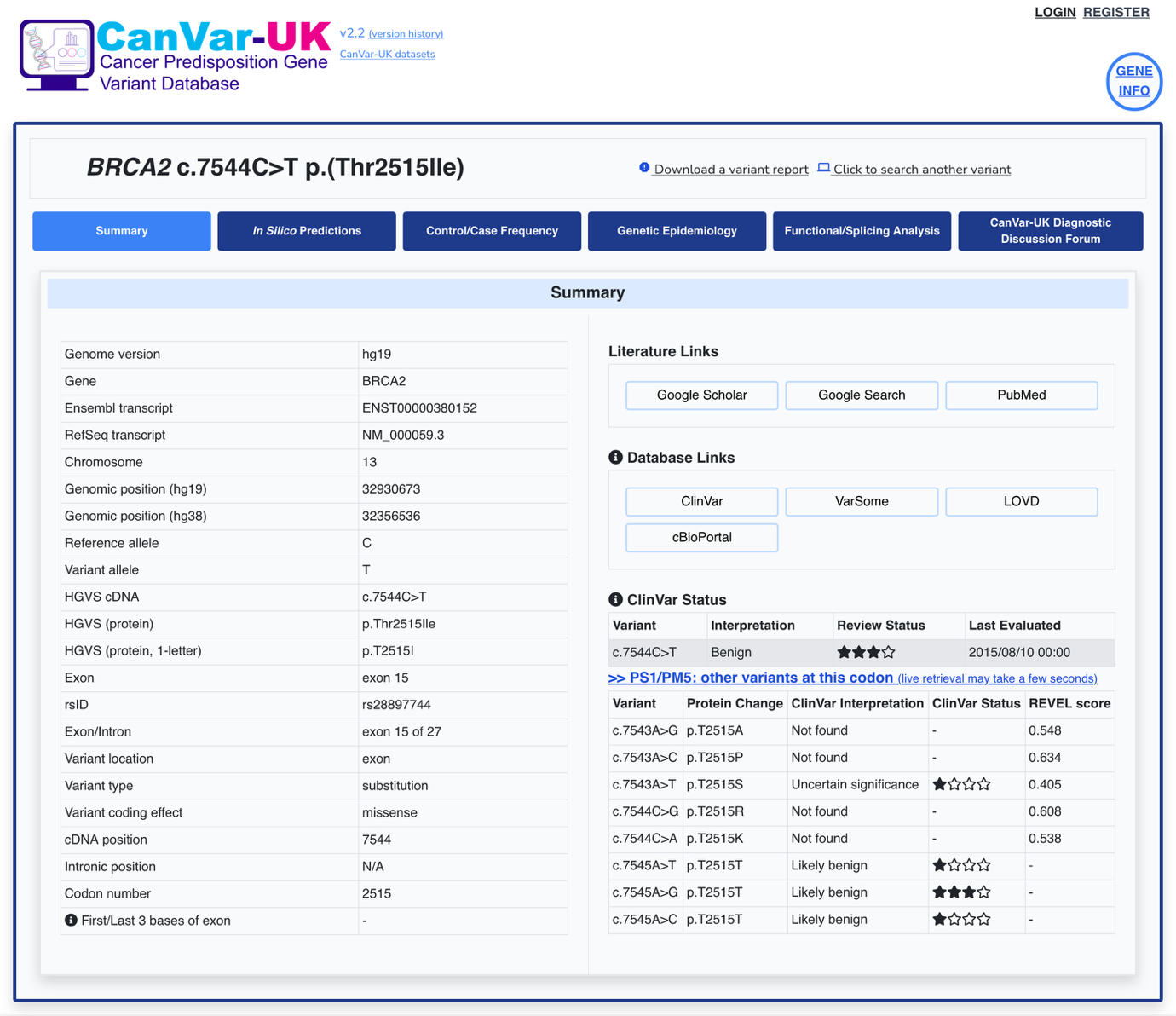

**Figure S1.** *CanVar-UK variant summary landing page for exemplar variant BRCA2 c.7544C>T.* Selection of a variant leads users to a landing page containing basic variant-specific information, including genomic position (in both hg19 and hg38), variant effect and both HGVSc and HGVSp. Links to external databases, including ClinVar, VarSome and cBioPortal are provided, and literature searches facilitated for PubMed, Google Scholar and standard Google searches via construction of variant-specific binary search strings. The ClinVar status of the variant is retrieved live using the ClinVar API, and a PS1/PM5 tool available to view classifications (alongside REVEL scores) for all other single nucleotide changes at the same codon.

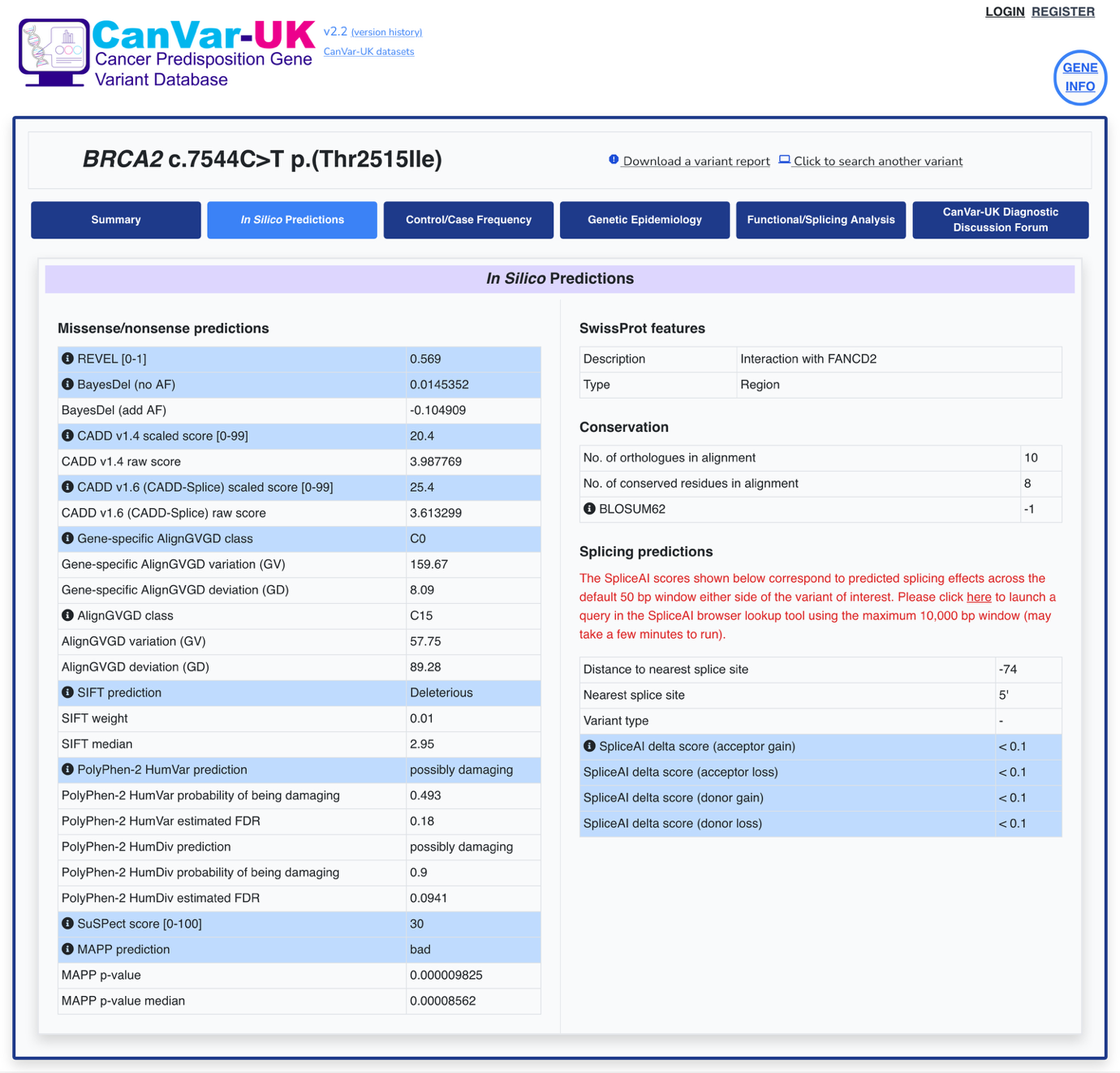

**Figure S2.** *CanVar-UK* in silico *prediction interface for exemplar variant BRCA2 c.7544C>T.* A selection of key *in silico* scores are collated for each variant, including REVEL, BayesDel and AlignGVGD. Information bubbles are presented alongside scores to aid in their interpretation. Domain information (via SwissProt) and conservation metrics are additionally displayed, as well as SpliceAI scores. A link is also generated allowing users to conduct a SpliceAI query for the variant using the maximum 10,000 bp splicing window in the SpliceAI browser lookup tool (<https://spliceailookup.broadinstitute.org/>).

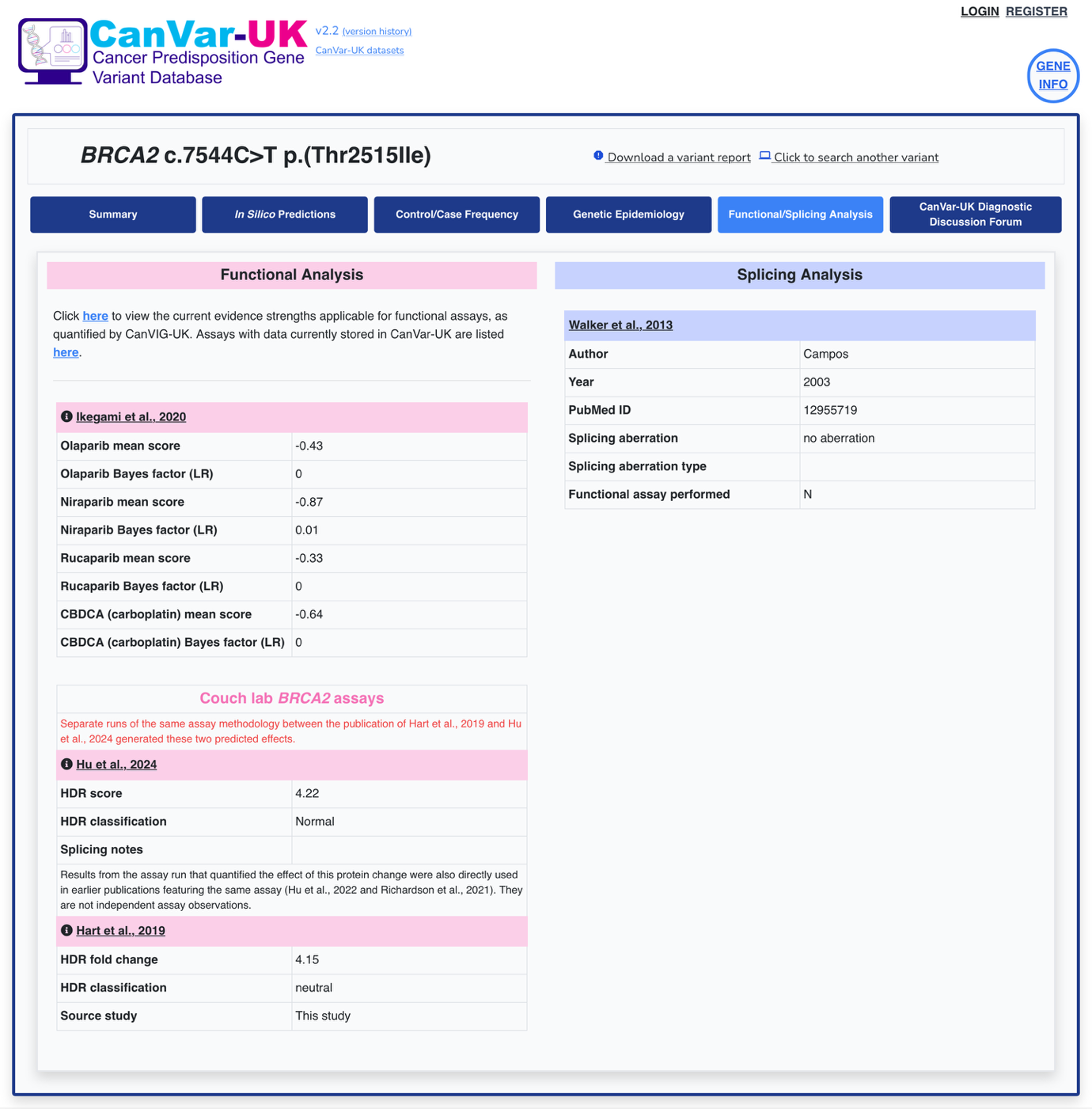

**Figure S3.** *CanVar-UK variant functional and splicing study interface for exemplar variant BRCA2 c.7544C>T.* CanVar-UK stores variant-level scores for 31 functional and splicing datasets encompassing 13 key cancer susceptibility genes (Table S2). Information bubbles accompany functional assay datasets to aid in interpretation of scores.

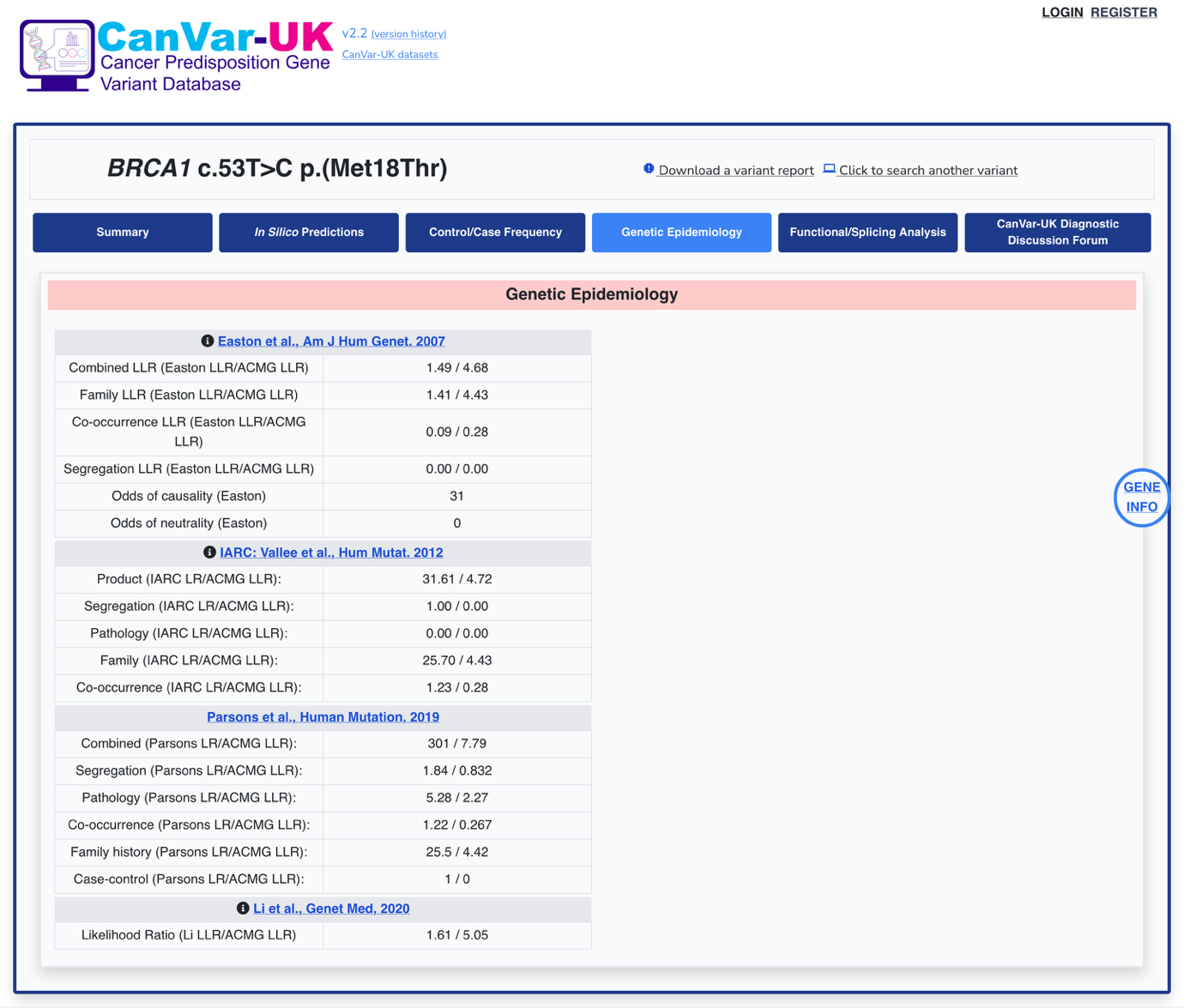

**Figure S4.** *CanVar-UK genetic epidemiology study interface page for exemplar variant BRCA1 c.53T>C.* Variant-level data from multifactorial genetic epidemiology studies in the *BRCA1* and *BRCA2* genes are collated and displayed, showing log-likelihood ratios (LLRs; including ACMG LLRs equivalent to exponent/evidence points) for both the combined and stratified epidemiological features.

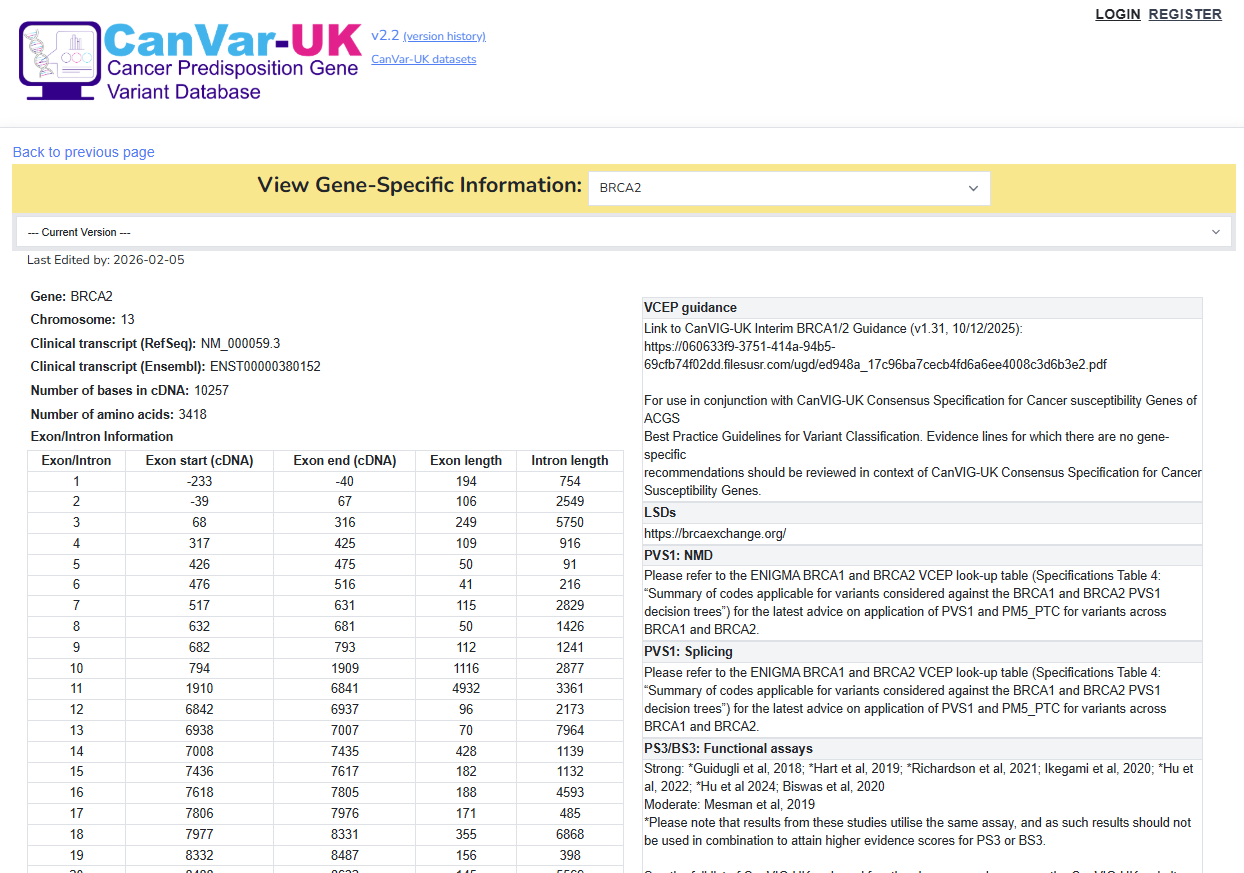

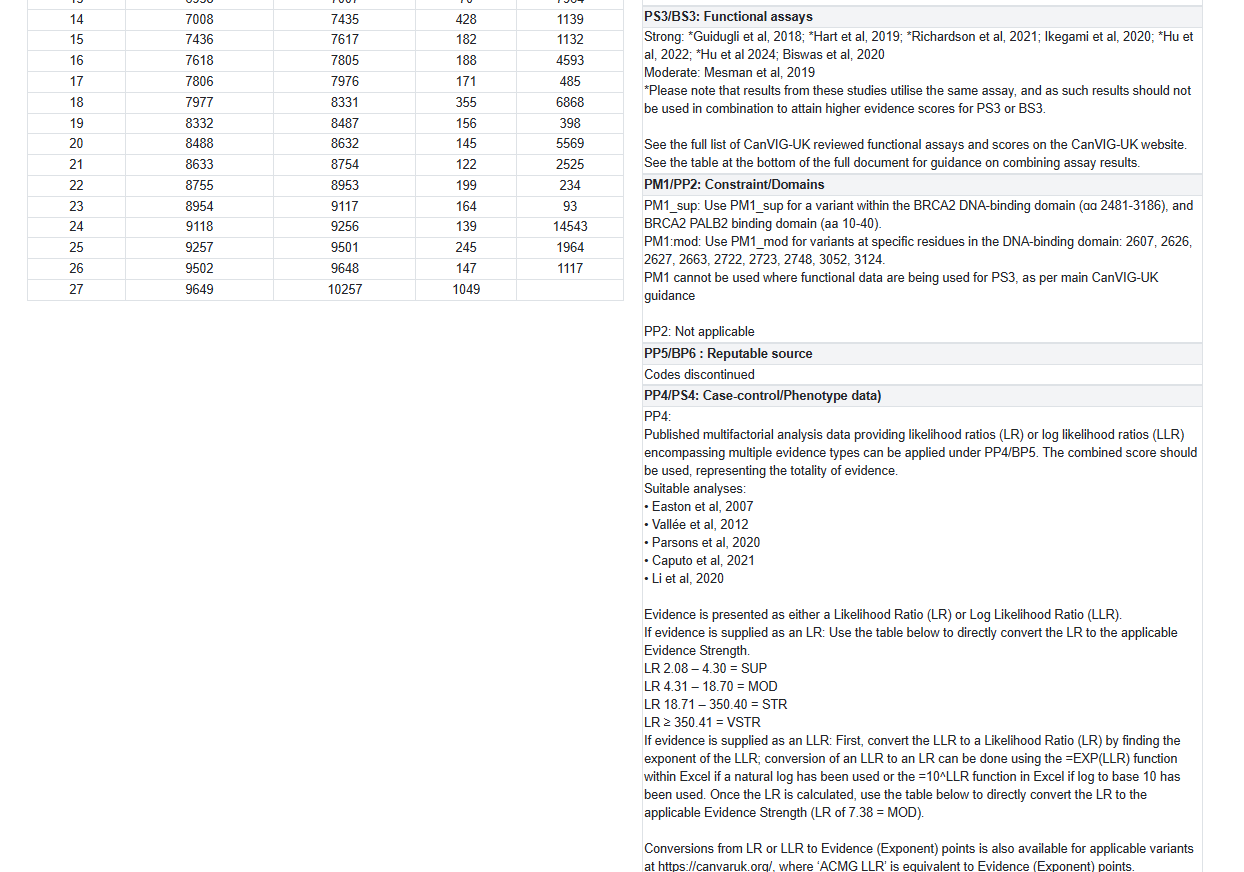

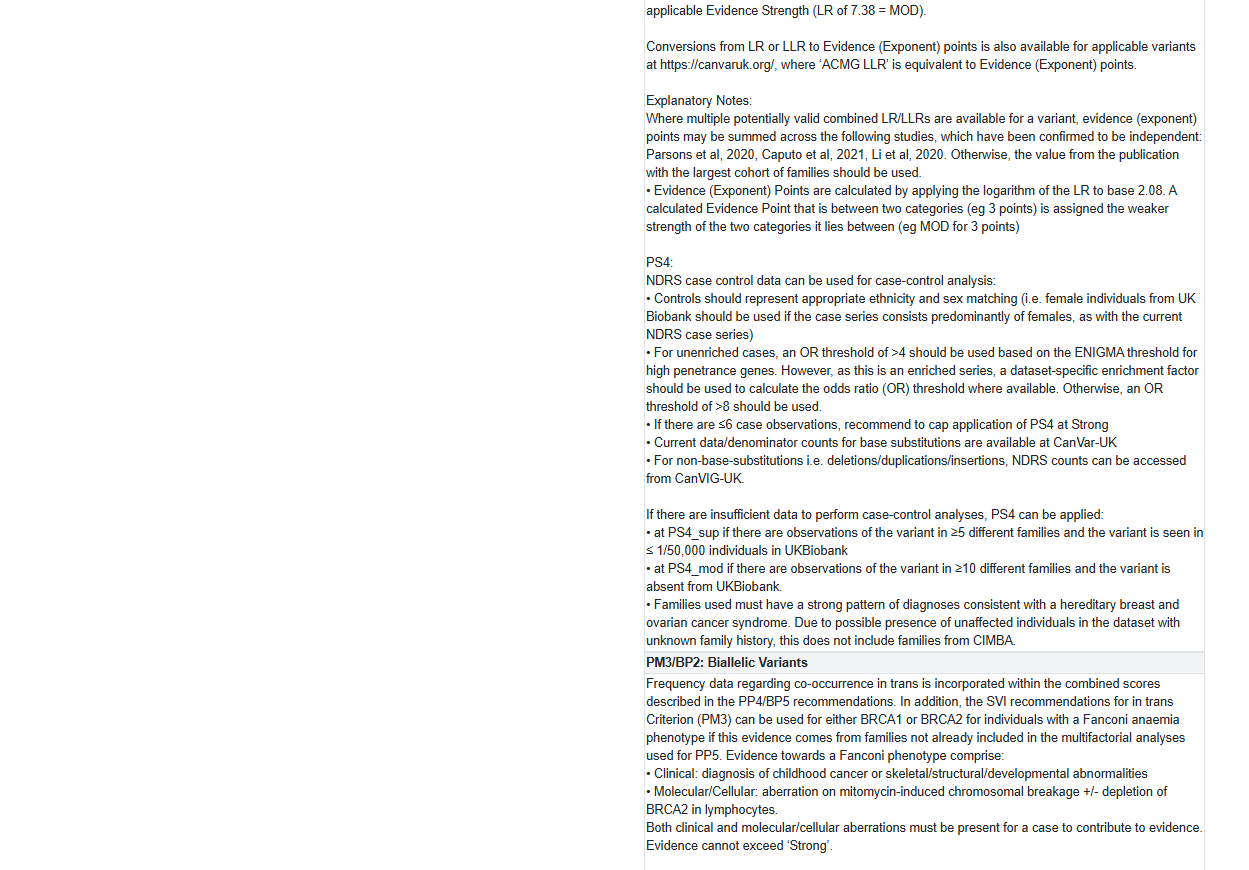

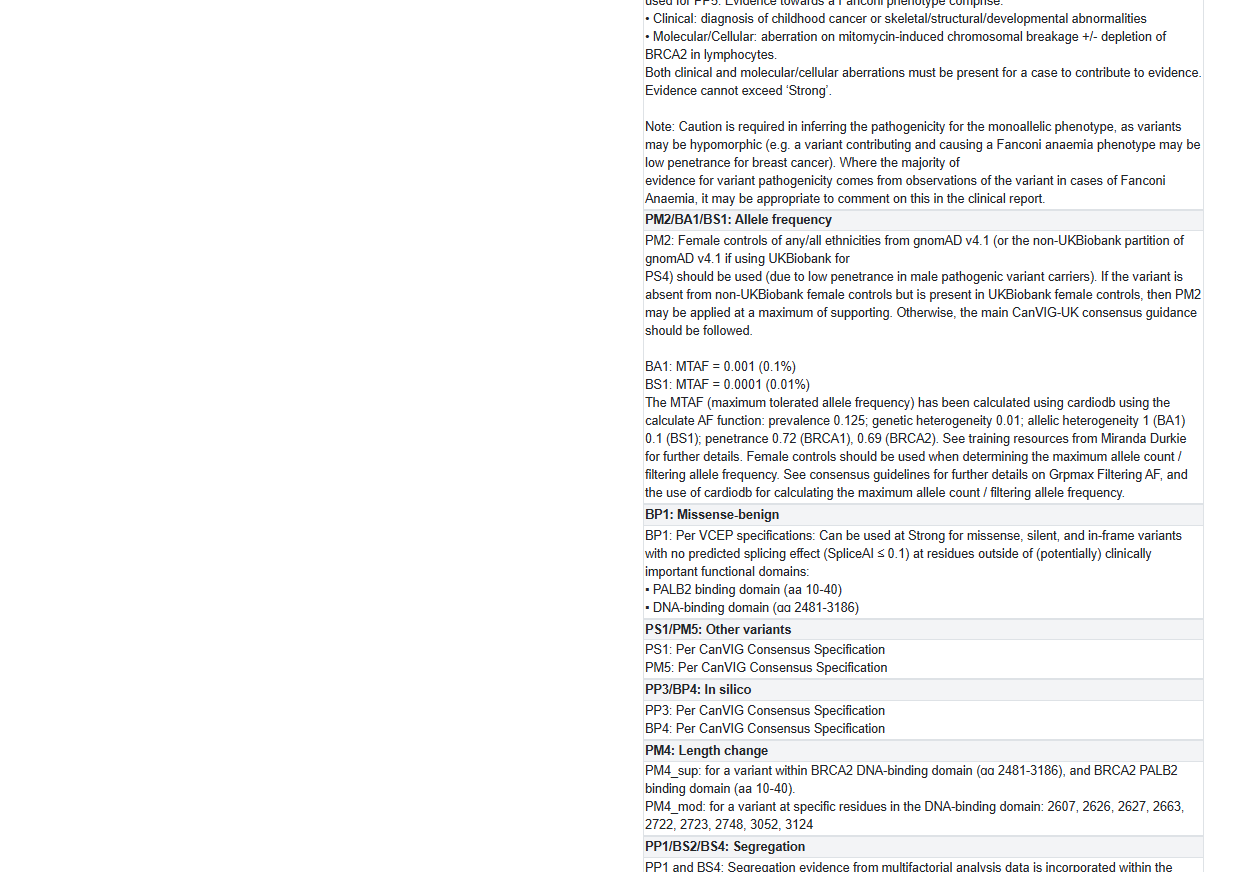

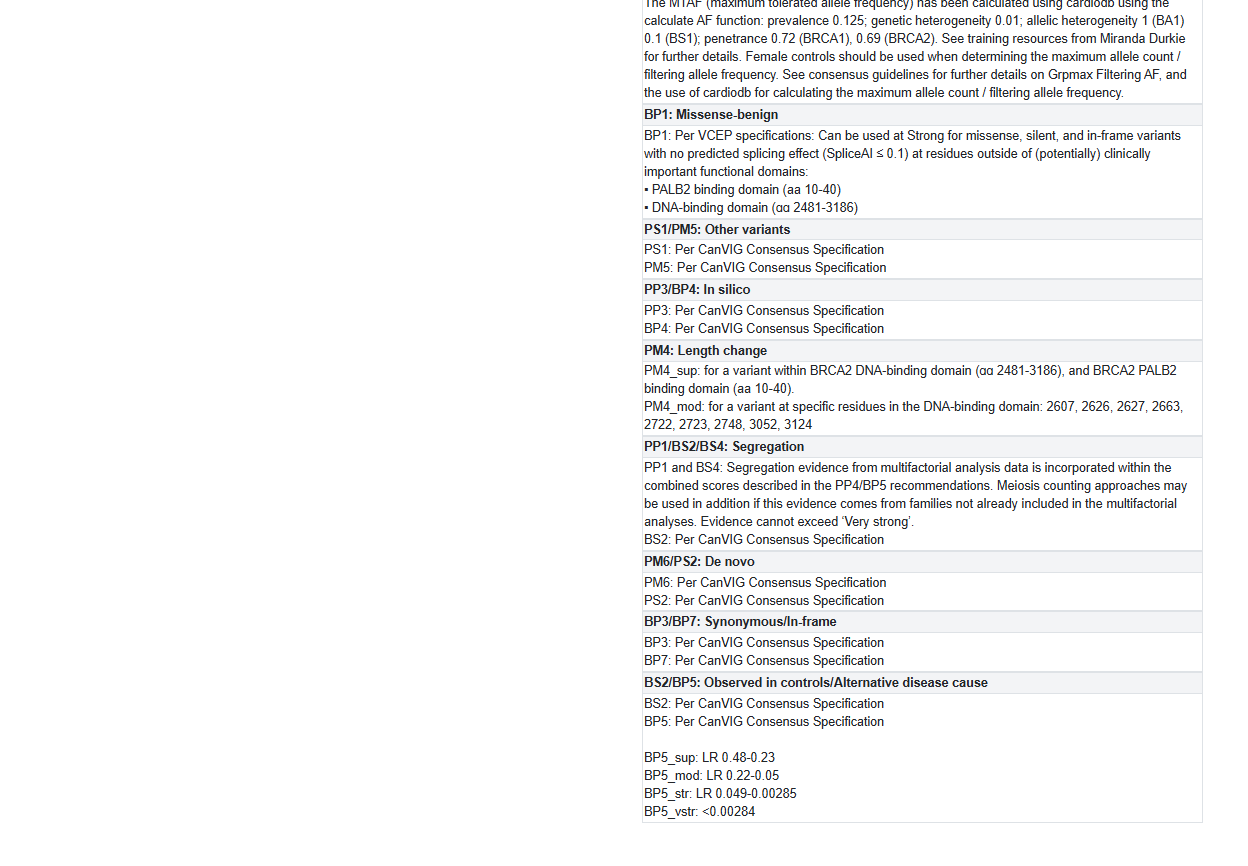

**Figure S5.** *Exemplar gene-specific information page for BRCA2.* Each gene listed in CanVar-UK has a gene-specific page that details basic features of the selected CanVar-UK transcript, such as cDNA and amino acid length, alongside the lengths and cDNA positions of its constituent exons and introns. For genes with VCEP guidance available, this is displayed, and is updated by the CanVar-UK team when new guidance versions are released. The gene-specific information can also be accessed from any variant-specific tab by clicking the “GENE INFO” bubble present on the upper right corner of the CarVar-UK interface, as illustrated in Figures S1-S4 and S6.

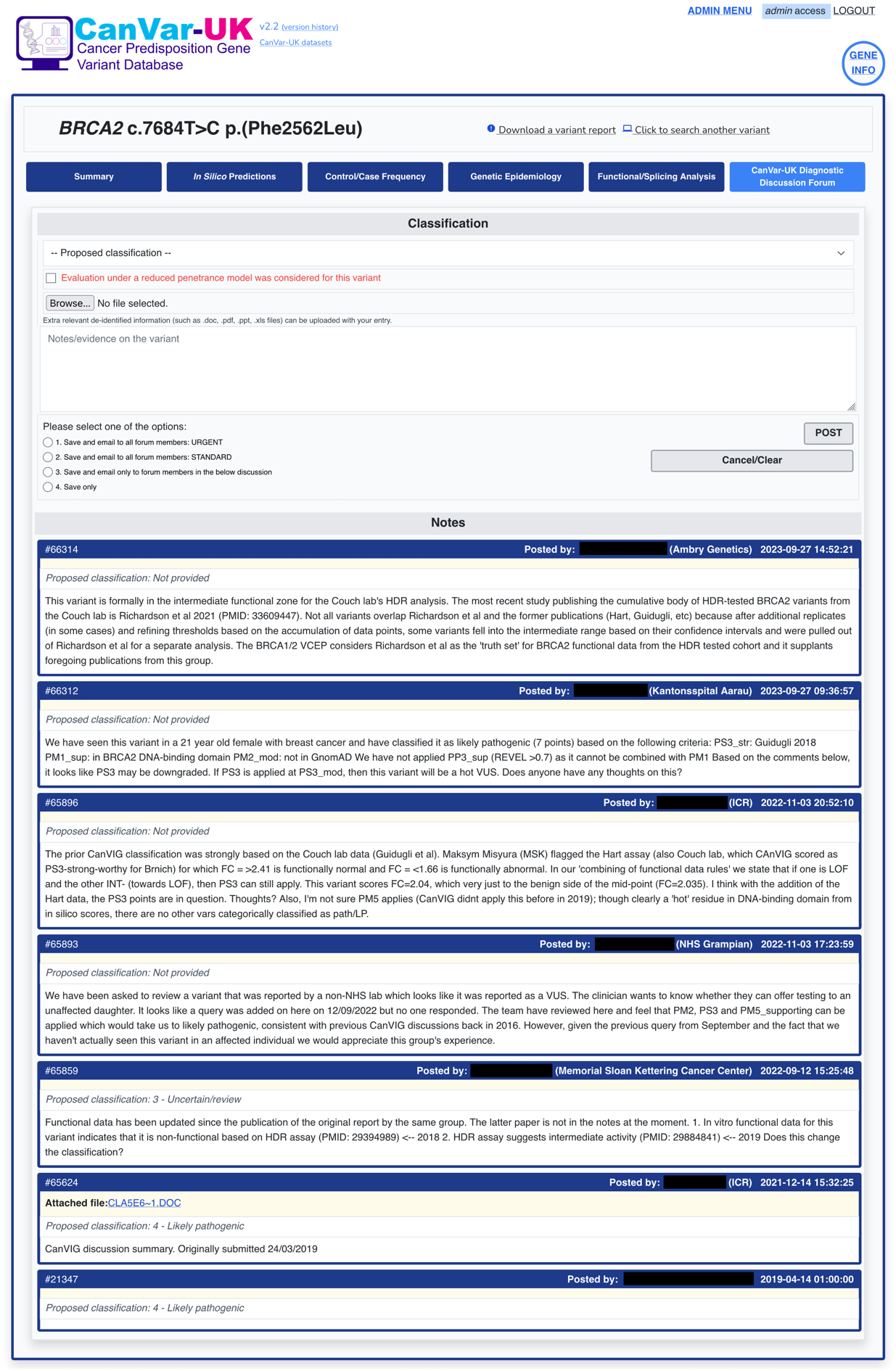

**Figure S6.** *CanVar-UK diagnostic discussion forum interface for exemplar variant BRCA2 c.7684C>T.* Users are able to submit posts to the CanVar-UK discussion forum for specific variants of interest, optionally with a proposed classification. These can either be simply saved for future reference, or can be sent out as an email marked “URGENT” or “STANDARD” to the CanVar user base, or to users who have previously posted about the variant.

**Figure S7.** *PDF variant report for exemplar variant BRCA2 c.7544C>T.* CanVar-UK offers a variant report functionality, using which a PDF report containing all key CanVar data fields associated with the variant can be generated. The final page of the report summarises key thresholds for evidence code application, and lists all functional studies that have been queried for the variant.

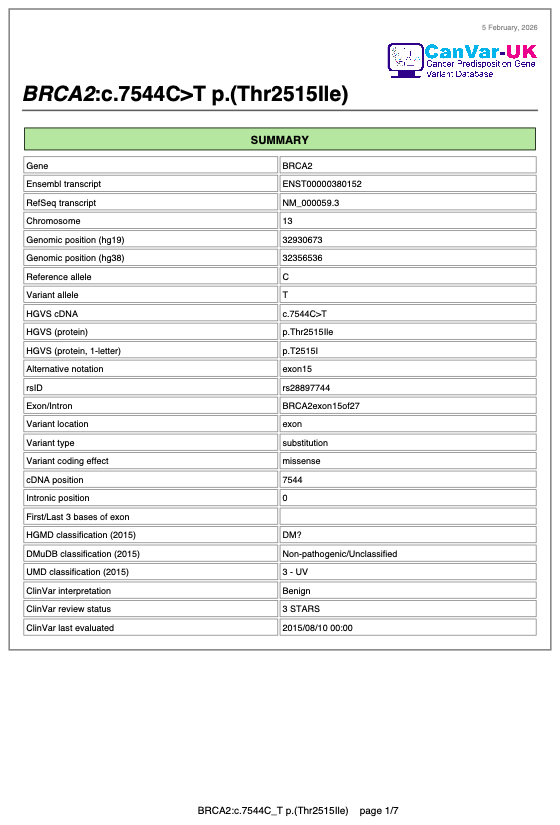

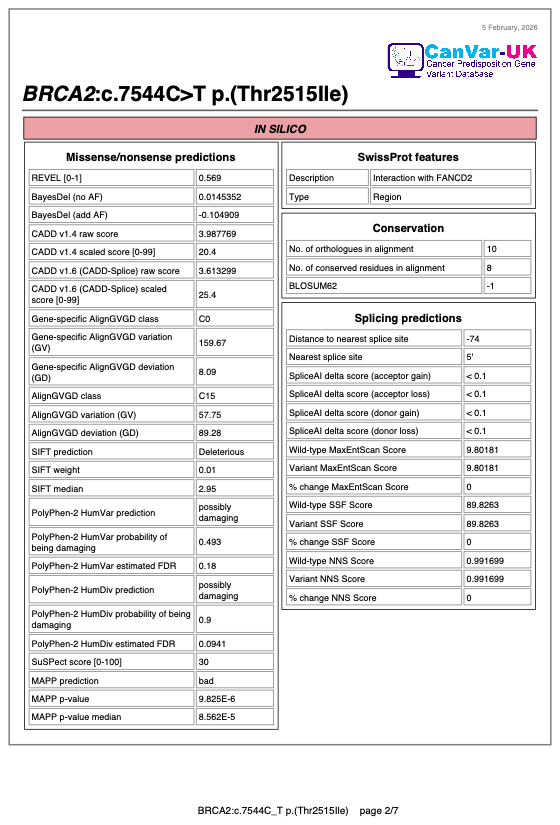

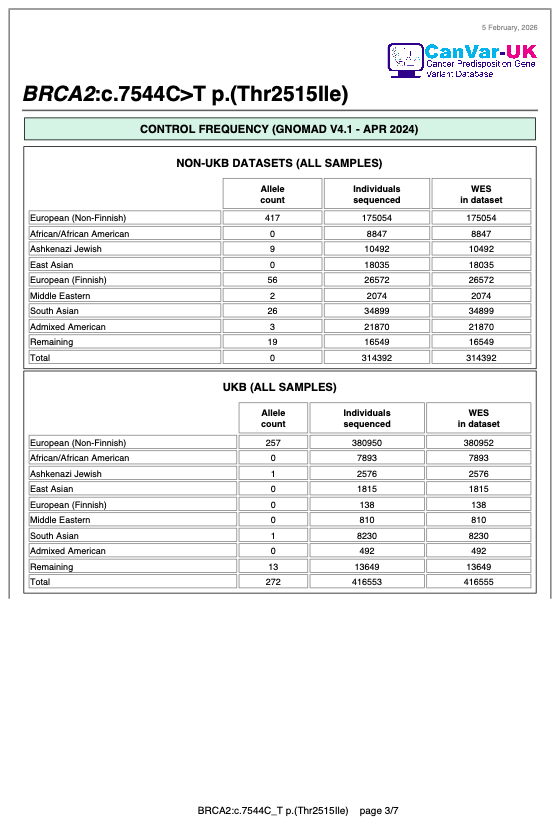

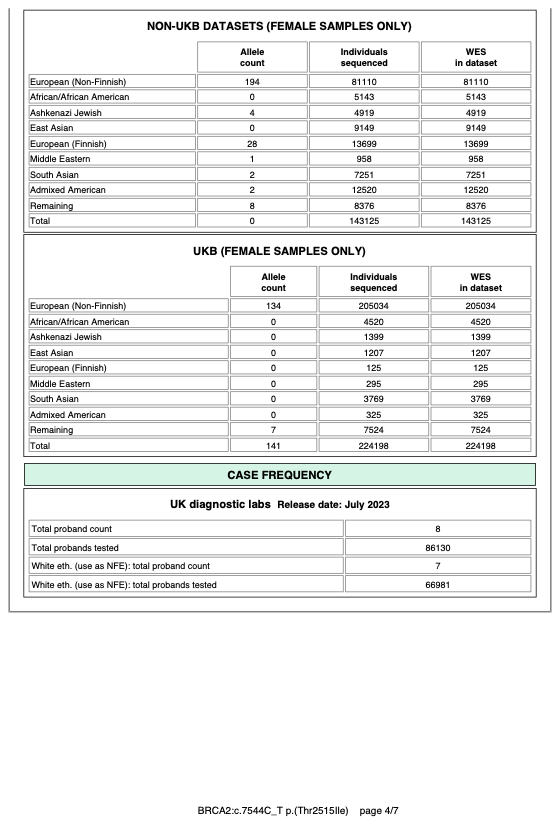

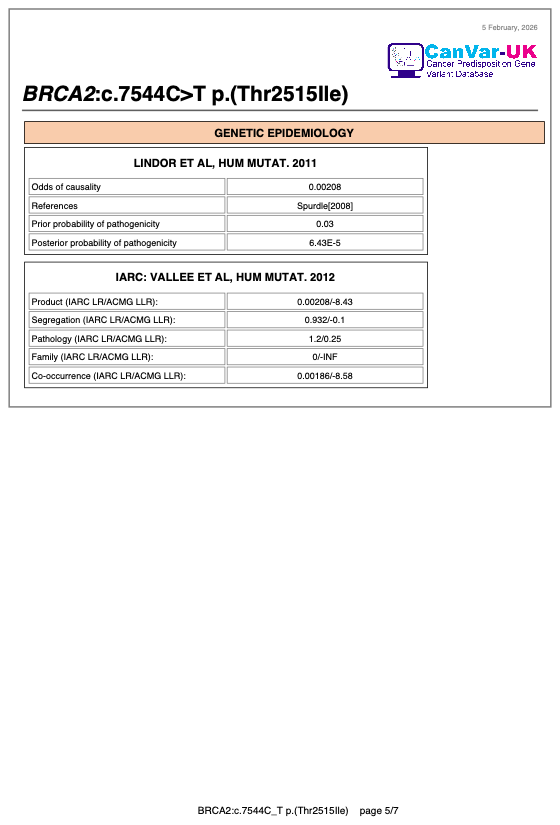

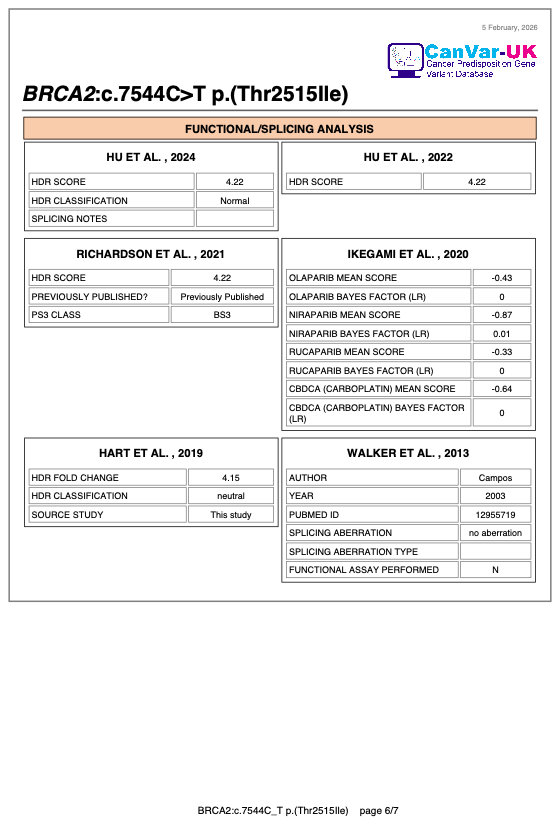

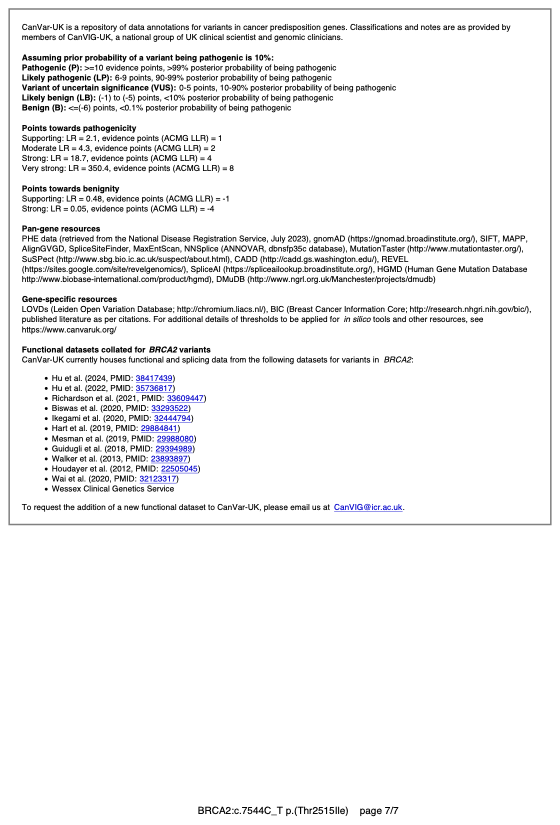

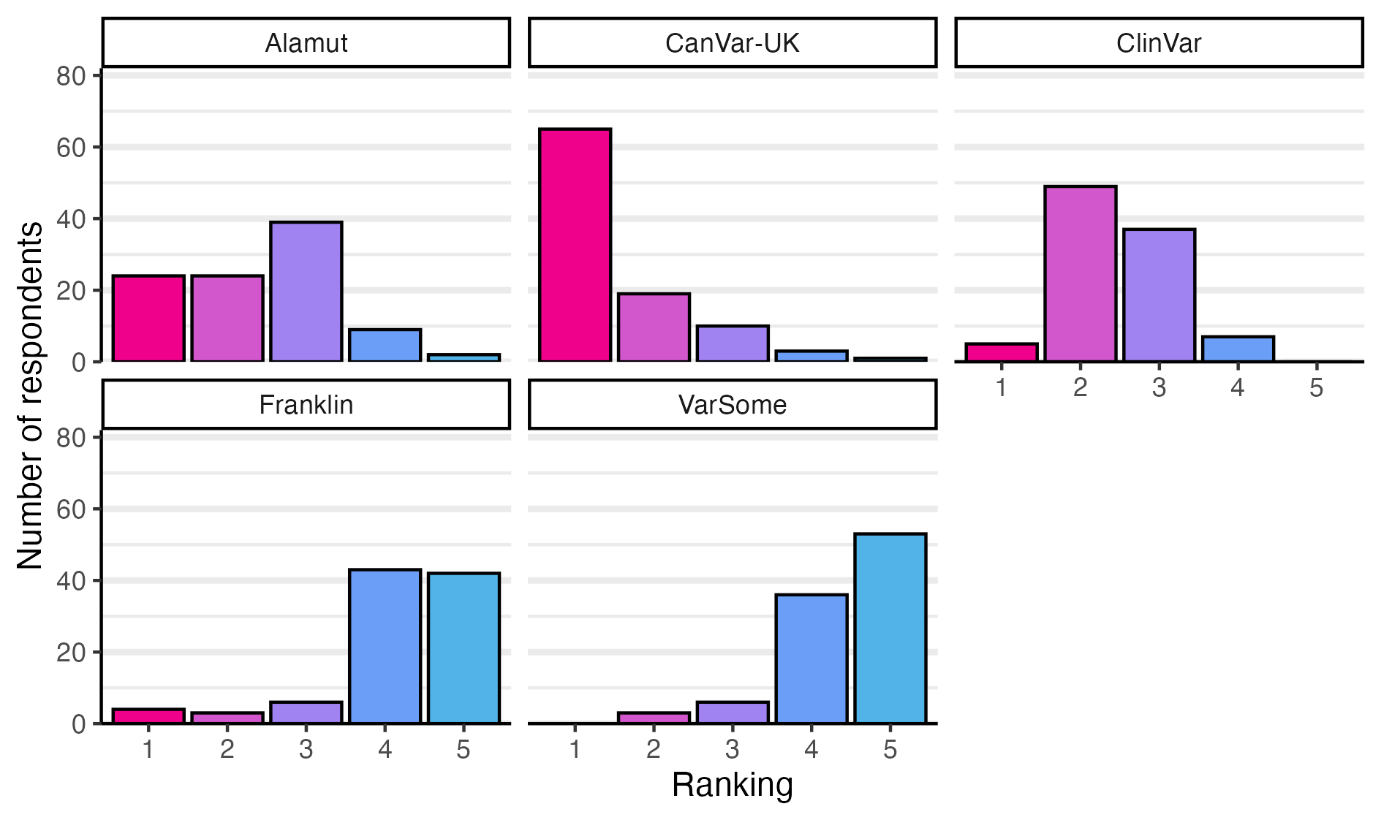

**Figure S8.** *Relative usefulness of variant interpretation platforms as ranked by respondents to the CanVIG-UK survey.* Shown are the responses to the prompt “Rank the following resources in order of how useful you find them for assembling evidence for variant interpretation in cancer susceptibility genes”, where a rank of 1 is the highest and 5 is the lowest. The responses displayed here are limited to clinical scientists (self-described role of “Qualified clinical scientist” or “Clinical scientist trainee”; the unfiltered counts of responses for all respondents are given in Table S3.

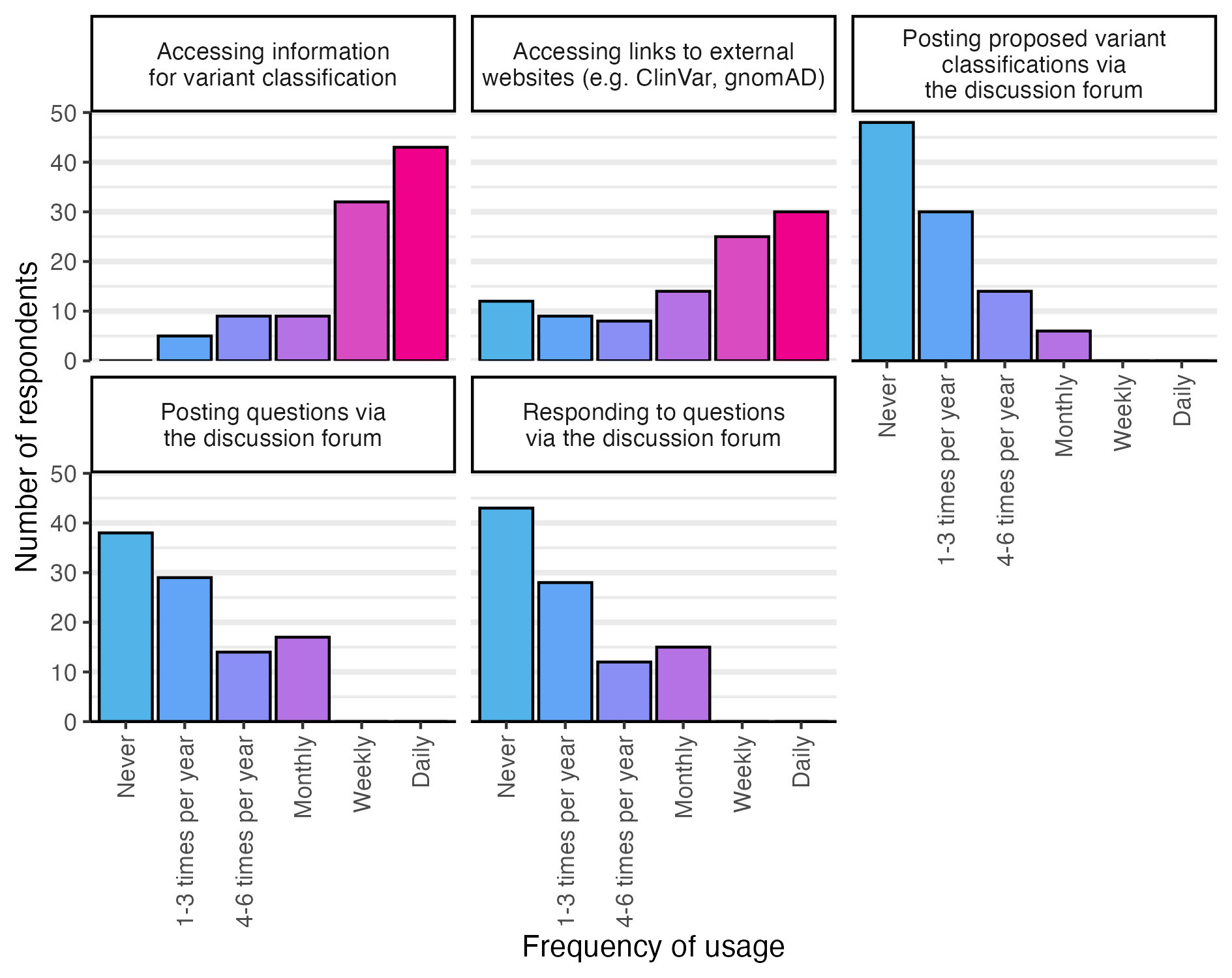

**Figure S9.** *CanVIG-UK survey responses evaluating frequency of CanVar-UK feature usage.* Shown are the responses to the question “How often on average do you use CanVar-UK for the following actions?” The responses displayed here are limited to clinical scientists (self-described role of “Qualified clinical scientist” or “Clinical scientist trainee”; the unfiltered counts of responses for all respondents are given in Table S3.

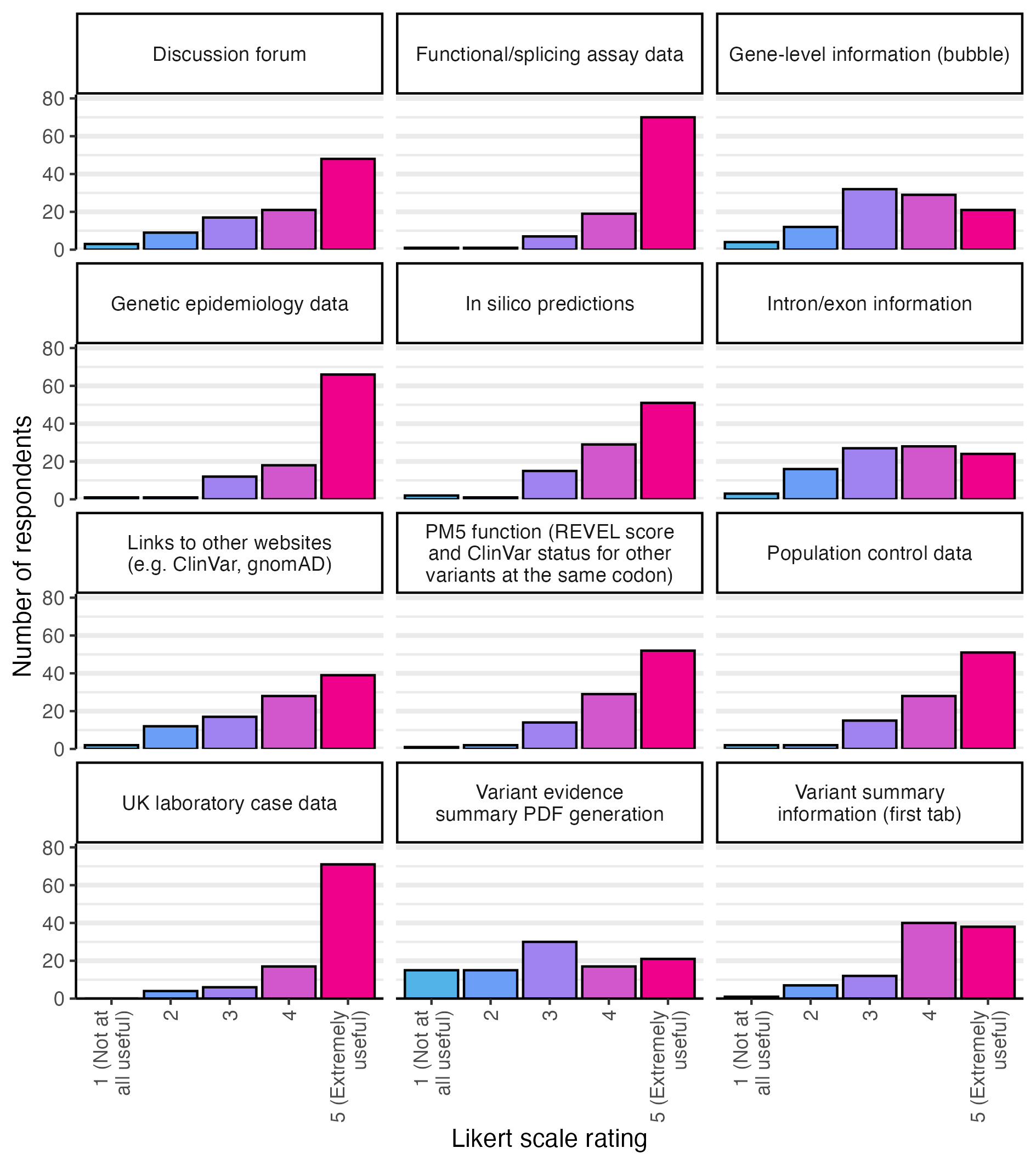

**Figure S10.** *Usefulness of individual CanVar-UK functionalities, as rated by CanVIG-UK members.* Shown are the responses to the question “How useful do you find the following CanVar-UK resources? (scale 1-5; 1=Not at all useful, 5=Extremely useful)”. The responses displayed here are limited to clinical scientists (self-described role of “Qualified clinical scientist” or “Clinical scientist trainee”; the unfiltered counts of responses for all respondents are given in Table S3.

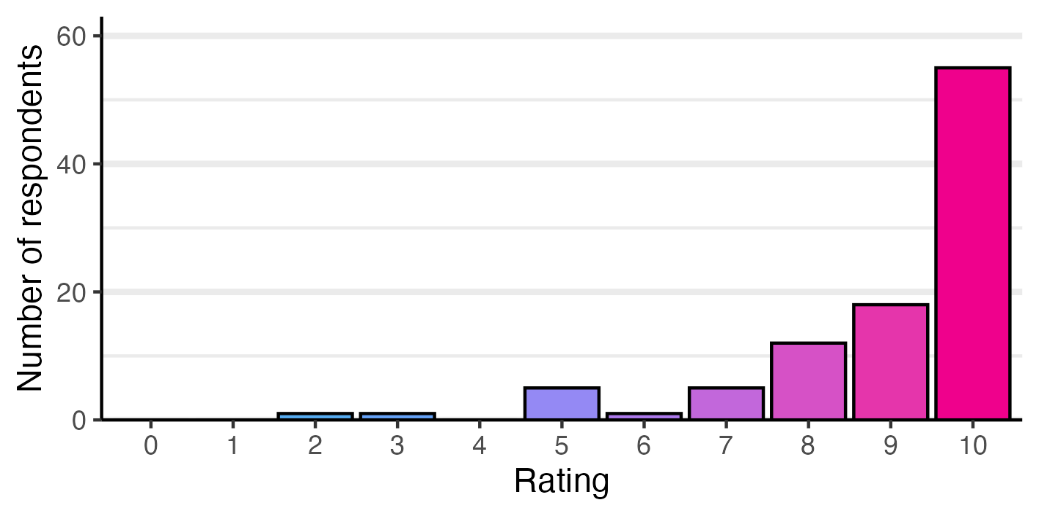

**Figure S11.** *Overall importance of the CanVar-UK platform, rated by CanVIG-UK members.* Shown are the responses to the question “Overall, how important is CanVar-UK for your work? (scale 0-10; 0=Not at all, 10=Essential)”. The responses displayed here are limited to clinical scientists (self-described role of “Qualified clinical scientist” or “Clinical scientist trainee”; the unfiltered counts of responses for all respondents are given in Table S3.

**Supplemental Tables**

**Table S1.** *List of CanVar-UK genes and transcripts with associated phenotypes.* Shown are the respective RefSeq and Ensembl transcripts for the 116 genes for which data is currently collated in ClinVar, alongside their respective (Online) Mendelian Inheritance in Man ((O)MIM) IDs and phenotypes and the names of any NHS germline cancer testing panels on which the gene is tested. Note that all OMIM phenotypes associated with a given gene are listed, and that not all of these may be associated with germline cancer susceptibility. AD, autosomal dominant; AR, autosomal recessive; NG, not given; XLR, X-linked recessive.

| **Gene** | **RefSeq transcript** | **Ensembl transcript** | **MIM ID** | **OMIM germline phenotype(s) (Inheritance pattern)** | **NHS panel (2025 genomic test directory)** |
| --- | --- | --- | --- | --- | --- |
| AIP | NM_003977.2 | ENST00000279146 | 605555 | Pituitary adenoma, multiple types (AD) Pituitary adenoma predisposition (AD) | Endocrine neoplasia |
| ALK | NM_004304.3 | ENST00000389048 | 105590 | Neuroblastoma, susceptibility to, 3 (NG) | Childhood solid tumours |
| ANKRD26 | NM_014915.2 | ENST00000376087 | 610855 | Thrombocytopenia 2 (AD) | Haematological malignancies cancer susceptibility; Inherited predisposition to acute myeloid leukaemia (AML) |
| APC | NM_000038.4 | ENST00000257430 | 611731 | Adenomatous polyposis coli (AD) Brain tumor-polyposis syndrome 2 (AD) Desmoid disease, hereditary (AD) Gardner syndrome (AD) Gastric adenocarcinoma and proximal polyposis of the stomach (AD) | Adult solid tumours cancer susceptibility; APC associated Polyposis; Brain cancer pertinent cancer susceptibility; Childhood solid tumours; Inherited polyposis and early onset colorectal cancer - germline testing |
| ATM | NM_000051.3 | ENST00000278616 | 607585 | Breast cancer, susceptibility to (AD) Ataxia-telangiectasia (AR) | Adult solid tumours cancer susceptibility; Brain cancer pertinent cancer susceptibility; Childhood solid tumours; Haematological malignancies cancer susceptibility; Inherited breast cancer and ovarian cancer; Inherited pancreatic cancer; Inherited prostate cancer |
| BAP1 | NM_004656.2 | ENST00000460680 | 603089 | Uveal melanoma, susceptibility to, 2 (AD) Kury-Isidor syndrome (AD) Tumor predisposition syndrome 1 (AD) | Adult solid tumours cancer susceptibility; BAP1 associated tumour predisposition syndrome; Childhood solid tumours; Familial melanoma; Inherited renal cancer |
| BARD1 | NM_000465.4 | ENST00000260947 | 601593 | Breast cancer, susceptibility to (AD) |  |
| BLM | NM_000057.2 | ENST00000355112 | 604610 | Bloom syndrome (AR) | Childhood solid tumours; Confirmed Fanconi anaemia or Bloom syndrome; Haematological malignancies cancer susceptibility |
| BMPR1A | NM_004329.2 | ENST00000372037 | 601299 | Polyposis syndrome, hereditary mixed, 2 (NG) Polyposis, juvenile intestinal (AD) | Adult solid tumours cancer susceptibility; Childhood solid tumours; Inherited polyposis and early onset colorectal cancer - germline testing |
| BRCA1 | NM_007294.3 | ENST00000357654 | 113705 | Breast-ovarian cancer, familial, 1 (AD) Pancreatic cancer, susceptibility to, 4 (NG) Fanconi anemia, complementation group S (AR) | Adult solid tumours cancer susceptibility; Breast cancer pertinent cancer susceptibility; Childhood solid tumours; Confirmed Fanconi anaemia or Bloom syndrome; Haematological malignancies cancer susceptibility; Inherited breast cancer and ovarian cancer; Inherited ovarian cancer (without breast cancer); Inherited pancreatic cancer; Inherited prostate cancer; Ovarian cancer pertinent cancer susceptibility |
| BRCA2 | NM_000059.3 | ENST00000380152 | 600185 | Breast cancer, male, susceptibility to (AD) Breast-ovarian cancer, familial, 2 (AD) Glioblastoma 3 (AR) Medulloblastoma (AD, AR) Pancreatic cancer 2 (NG) Prostate cancer (AD) Fanconi anemia, complementation group D1 (AR) Wilms tumor (AD) | Adult solid tumours cancer susceptibility; Breast cancer pertinent cancer susceptibility; Childhood solid tumours; Confirmed Fanconi anaemia or Bloom syndrome; Haematological malignancies cancer susceptibility; Inherited breast cancer and ovarian cancer; Inherited ovarian cancer (without breast cancer); Inherited pancreatic cancer; Inherited prostate cancer; Ovarian cancer pertinent cancer susceptibility |
| BRIP1 | NM_032043.2 | ENST00000259008 | 605882 | Breast cancer, early-onset, susceptibility to (AD) Fanconi anemia, complementation group J (NG) | Adult solid tumours cancer susceptibility; Childhood solid tumours; Confirmed Fanconi anaemia or Bloom syndrome; Haematological malignancies cancer susceptibility; Inherited ovarian cancer (without breast cancer); Ovarian cancer pertinent cancer susceptibility |
| BUB1B | NM_001211.5 | ENST00000287598 | 602860 | Premature chromatid separation trait (AD) Mosaic variegated aneuploidy syndrome (AR) | Childhood solid tumours |
| CBL | NM_005188.3 | ENST00000264033 | 165360 | ?Juvenile myelomonocytic leukemia (AD) Noonan syndrome-like disorder with or without juvenile myelomonocytic leukemia (AD) | Adult solid tumours cancer susceptibility; Childhood solid tumours; Haematological malignancies cancer susceptibility |
| CDC73 | NM_024529.4 | ENST00000367435 | 607393 | Hyperparathyroidism-jaw tumor syndrome (AD) Parathyroid adenoma with cystic changes (AD) | Adult solid tumours cancer susceptibility; Childhood solid tumours; Endocrine neoplasia; Inherited parathyroid cancer |
| CDH1 | NM_004360.3 | ENST00000261769 | 192090 | Blepharocheilodontic syndrome 1 (AD) Diffuse gastric and lobular breast cancer syndrome with or without cleft lip and/or palate (AD) | Adult solid tumours cancer susceptibility; Hereditary diffuse gastric cancer |
| CDK4 | NM_000075.2 | ENST00000257904 | 123829 | Melanoma, cutaneous malignant, 3 (AD) | Adult solid tumours cancer susceptibility; Familial melanoma |
| CDKN1C | NM_000076.2 | ENST00000414822 | 130650 | Beckwith-Wiedemann syndrome (AD) IMAGE syndrome (AD) | Childhood solid tumours; Wilms tumour with features suggestive of predisposition |
| CDKN2A | NM_000077.4 | ENST00000304494 | 600160 | Melanoma and neural system tumor syndrome (AD) Melanoma-pancreatic cancer syndrome (AD) Melanoma, cutaneous malignant, 2 (AD) | Adult solid tumours cancer susceptibility; Childhood solid tumours; Familial melanoma; Inherited pancreatic cancer |
| CEBPA | NM_004364.3 | ENST00000498907 | 116897 | ?Leukemia, acute myeloid (AD) | Haematological malignancies cancer susceptibility; Inherited predisposition to acute myeloid leukaemia (AML) |
| CEP57 | NM_014679.3 | ENST00000325542 | 607951 | Mosaic variegated aneuploidy syndrome (AR) |  |
| CHEK2 | NM_007194.3 | ENST00000328354 | 604373 | Tumor predisposition syndrome 4, breast / prostate / colorectal (NG) | Inherited breast cancer and ovarian cancer; Inherited prostate cancer |
| CYLD | NM_015247.2 | ENST00000311559 | 605018 | ?Frontotemporal dementia and / or amyotrophic lateral sclerosis (AD) Brooke-Spiegler syndrome (AD) Cylindromatosis, familial (AD) Trichoepithelioma, multiple familial, 1 (AD) | Multiple monogenic benign skin tumours |
| DDB2 | NM_000107.2 | ENST00000256996 | 600811 | Xeroderma pigmentosum, group E, DDB-negative subtype (AR) | Adult solid tumours cancer susceptibility; Childhood solid tumours; Xeroderma pigmentosum, Trichothiodystrophy or Cockayne syndrome |
| DDX41 | NM_016222.3 | ENST00000507955 | 608170 | Myeloproliferative / lymphoproliferative neoplasms, familial (multiple types), susceptibility to (AD) | Haematological malignancies cancer susceptibility; Inherited predisposition to acute myeloid leukaemia (AML) |
| DICER1 | NM_177438.2 | ENST00000343455 | 138800 | Goiter, multinodular 1, with or without Sertoli-Leydig cell tumors (AD) Pleuropulmonary blastoma (AD) Rhabdomyosarcoma, embryonal, 2 (NG) | Adult solid tumours cancer susceptibility; Childhood solid tumours; DICER1-related cancer predisposition |
| DIS3L2 | NM_152383.4 | ENST00000325385 | 614184 | Perlman syndrome (AR) | Childhood solid tumours |
| EGFR | NM_005228.3 | ENST00000275493 | 131550 | Nonsmall cell lung cancer, susceptibility to (AD) Adenocarcinoma of lung, response to tyrosine kinase inhibitor in (AD) Neonatal nephrocutaneous inflammatory syndrome (AR) Nonsmall cell lung cancer, response to tyrosine kinase inhibitor in (AD) |  |
| EPCAM | NM_002354.2 | ENST00000263735 | 185535 | Diarrhea, 5, with tufting enteropathy, congenital (AR) Lynch syndrome 8 (AR) | Adult solid tumours cancer susceptibility; Inherited MMR deficiency (Lynch syndrome); Inherited polyposis and early onset colorectal cancer - germline testing |
| ERCC2 | NM_000400.3 | ENST00000391945 | 126340 | ?Cerebrooculofacioskeletal syndrome 2 (AR) Trichothiodystrophy 1, photosensitive (AR) Xeroderma pigmentosum, group D (AR) | Adult solid tumours cancer susceptibility; Childhood solid tumours |
| ERCC3 | NM_000122.1 | ENST00000285398 | 133510 | Trichothiodystrophy 2, photosensitive (AR) Xeroderma pigmentosum, group B (AR) | Adult solid tumours cancer susceptibility; Childhood solid tumours |
| ERCC4 | NM_005236.2 | ENST00000311895 | 133520 | Fanconi anemia, complementation group Q (AR) Xeroderma pigmentosum, type F / Cockayne syndrome (AR) XFE progeroid syndrome (AR) | Adult solid tumours cancer susceptibility; Childhood solid tumours; Confirmed Fanconi anaemia or Bloom syndrome; Haematological malignancies cancer susceptibility |
| ERCC5 | NM_000123.2 | ENST00000355739 | 133530 | Cerebrooculofacioskeletal syndrome 3 (AR) Xeroderma pigmentosum, group G / Cockayne syndrome (AR) | Adult solid tumours cancer susceptibility; Childhood solid tumours |
| ETV6 | NM_001987.4 | ENST00000396373 | 600618 | Thrombocytopenia 5 (AD) | Haematological malignancies cancer susceptibility; Inherited predisposition to acute myeloid leukaemia (AML); Inherited susceptibility to acute lymphoblastoid leukaemia (ALL) |
| EXT1 | NM_000127.2 | ENST00000378204 | 133700 | Exostoses, multiple, type 1 (AD) | Multiple exostoses; Sarcoma susceptibility |
| EXT2 | NM_000401.3 | ENST00000395673 | 608210 | Exostoses, multiple, type 2 (AD) Seizures, scoliosis, and macrocephaly syndrome (AR) | Multiple exostoses; Sarcoma susceptibility |
| EZH2 | NM_004456.4 | ENST00000320356 | 601573 | Weaver syndrome (AD) | Childhood solid tumours |
| FANCA | NM_000135.2 | ENST00000389301 | 227650 | Fanconi anemia, complementation group A (AR) | Adult solid tumours cancer susceptibility; Childhood solid tumours; Confirmed Fanconi anaemia or Bloom syndrome; Haematological malignancies cancer susceptibility |
| FANCB | NM_001018113.1 | ENST00000398334 | 300515 | Fanconi anemia, complementation group B (XLR) | Adult solid tumours cancer susceptibility; Childhood solid tumours; Confirmed Fanconi anaemia or Bloom syndrome; Haematological malignancies cancer susceptibility |
| FANCC | NM_000136.2 | ENST00000289081 | 613899 | Fanconi anemia, complementation group C (AR) | Adult solid tumours cancer susceptibility; Childhood solid tumours; Confirmed Fanconi anaemia or Bloom syndrome; Haematological malignancies cancer susceptibility |
| FANCD2 | NM_033084.3 | ENST00000287647 | 613984 | Fanconi anemia, complementation group D2 (AR) | Adult solid tumours cancer susceptibility; Childhood solid tumours; Confirmed Fanconi anaemia or Bloom syndrome; Haematological malignancies cancer susceptibility |
| FANCE | NM_021922.2 | ENST00000229769 | 613976 | Fanconi anemia, complementation group E (AR) | Adult solid tumours cancer susceptibility; Childhood solid tumours; Confirmed Fanconi anaemia or Bloom syndrome; Haematological malignancies cancer susceptibility |
| FANCF | NM_022725.3 | ENST00000327470 | 613897 | Fanconi anemia, complementation group F (AR) | Adult solid tumours cancer susceptibility; Childhood solid tumours; Confirmed Fanconi anaemia or Bloom syndrome; Haematological malignancies cancer susceptibility |
| FANCG | NM_004629.1 | ENST00000378643 | 602956 | Fanconi anemia, complementation group G (AR) | Adult solid tumours cancer susceptibility; Childhood solid tumours; Confirmed Fanconi anaemia or Bloom syndrome; Haematological malignancies cancer susceptibility |
| FANCI | NM_001113378.1 | ENST00000310775 | 611360 | Fanconi anemia, complementation group I (AR) | Adult solid tumours cancer susceptibility; Childhood solid tumours; Confirmed Fanconi anaemia or Bloom syndrome; Haematological malignancies cancer susceptibility |
| FANCL | NM_001114636.1 | ENST00000402135 | 608111 | Fanconi anemia, complementation group L (AR) | Adult solid tumours cancer susceptibility; Childhood solid tumours; Confirmed Fanconi anaemia or Bloom syndrome; Haematological malignancies cancer susceptibility |
| FANCM | NM_020937.2 | ENST00000267430 | 609644 | Premature ovarian failure (AR) Spermatogenic failure 28 (AR) |  |
| FH | NM_000143.3 | ENST00000366560 | 136850 | Fumarase deficiency (AR) Leiomyomatosis and renal cell cancer (AD) | Adult solid tumours cancer susceptibility; Inherited phaeochromocytoma and paraganglioma excluding NF1; Inherited renal cancer |
| FLCN | NM_144997.5 | ENST00000285071 | 135150 | Birt-Hogg-Dube syndrome (AD) | Adult solid tumours cancer susceptibility; Inherited renal cancer; Multiple monogenic benign skin tumours |
| GATA1 | NM_002049.4 | ENST00000376670 | 305371 | Anemia, congenital, nonspherocytic hemolytic, 9 (XLR) Anemia, X-linked, with / without neutropenia and / or platelet abnormalities (XLR) Thrombocytopenia with beta-thalassemia, X-linked (XLR) Thrombocytopenia, X-linked, with or without dyserythropoietic anemia (XLR) | Haematological malignancies cancer susceptibility |
| GATA2 | NM_032638.4 | ENST00000341105 | 137295 | Leukemia, acute myeloid, susceptibility to (AD) Myelodysplastic syndrome, susceptibility to (NG) Emberger syndrome (AD) Immunodeficiency (AD) | Haematological malignancies cancer susceptibility; Inherited predisposition to acute myeloid leukaemia (AML) |
| GPC3 | NM_004484.3 | ENST00000370818 | 300037 | Simpson-Golabi-Behmel syndrome, type 1 (XLR) | Childhood solid tumours |
| GREM1 | NM_013372.6 | ENST00000300177 | 603054 | Polyposis syndrome, hereditary mixed, 1 (AD) | Inherited polyposis and early onset colorectal cancer - germline testing |
| HNF1A | NM_000545.5 | ENST00000257555 | 142410 | Diabetes mellitus, insulin dependent (AR) Diabetes mellitus, noninsulin-dependent, 2 (AD) Diabetes mellitus, insulin-dependent, 20 (NG) MODY, type III (AD) Renal cell carcinoma (NG) |  |
| HRAS | NM_005343.2 | ENST00000311189 | 190020 | Congenital myopathy with excess of muscle spindles / Costello syndrome (AD) | Adult solid tumours cancer susceptibility; Childhood solid tumours |
| KIT | NM_000222.2 | ENST00000288135 | 164290 | Gastrointestinal stromal tumor, familial (AD) Mastocytosis, cutaneous (AD) Piebaldism (AD) | Adult solid tumours cancer susceptibility; Inherited predisposition to GIST |
| KRAS | NM_004985.4 | ENST00000311936 | 190070 | Cardiofaciocutaneous syndrome 2 (AD) Noonan syndrome 3 (AD) RAS-associated autoimmune leukoproliferative disorder (AD) | Adult solid tumours cancer susceptibility; Childhood solid tumours |
| LZTR1 | NM_006767.3 | ENST00000215739 | 600574 | Schwannomatosis-2, susceptibility to (AD) Noonan syndrome 10 (AD) Noonan syndrome 2 (AR) | Familial tumours of the nervous system |
| MAX | NM_002382.3 | ENST00000358664 | 154950 | Pheochromocytoma, susceptibility to (AD) Polydactyly-macrocephaly syndrome (AD) | Adult solid tumours cancer susceptibility; Childhood solid tumours; Inherited phaeochromocytoma and paraganglioma excluding NF1 |
| MEN1 | NM_000244.3 | ENST00000443283 | 613733 | Multiple endocrine neoplasia 1 (AD) | Adult solid tumours cancer susceptibility; Childhood solid tumours; Endocrine neoplasia; Inherited phaeochromocytoma and paraganglioma excluding NF1 |
| MET | NM_001127500.1 | ENST00000318493 | 164860 | ?Arthrogryposis, distal, type 11 (AD) ?Deafness, autosomal recessive 97 (AR) Osteofibrous dysplasia, susceptibility to (AD) Renal cell carcinoma, papillary, 1, familial and somatic (NG) | Adult solid tumours cancer susceptibility; Inherited renal cancer |
| MITF | NM_000248.3 | ENST00000394351 | 156845 | Melanoma, cutaneous malignant, susceptibility to, 8 (NG) COMMAD syndrome (AR) Tietz albinism-deafness syndrome (AD) Waardenburg syndrome, type 2A (AD) |  |
| MLH1 | NM_000249.3 | ENST00000231790 | 120436 | Lynch syndrome 2 (NG) Mismatch repair cancer syndrome 1 (AR) Muir-Torre syndrome (AD) | Adult solid tumours cancer susceptibility; Brain cancer pertinent cancer susceptibility; Childhood solid tumours; Haematological malignancies cancer susceptibility; Inherited MMR deficiency (Lynch syndrome); Inherited ovarian cancer (without breast cancer); Inherited polyposis and early onset colorectal cancer - germline testing; Inherited prostate cancer; Multiple monogenic benign skin tumours; Ovarian cancer pertinent cancer susceptibility |
| MSH2 | NM_000251.1 | ENST00000233146 | 609309 | Lynch syndrome 1 (AD) Mismatch repair cancer syndrome 2 (AR) Muir-Torre syndrome (AD) | Adult solid tumours cancer susceptibility; Brain cancer pertinent cancer susceptibility; Childhood solid tumours; Haematological malignancies cancer susceptibility; Inherited MMR deficiency (Lynch syndrome); Inherited ovarian cancer (without breast cancer); Inherited polyposis and early onset colorectal cancer - germline testing; Inherited prostate cancer; Multiple monogenic benign skin tumours; Ovarian cancer pertinent cancer susceptibility |
| MSH6 | NM_000179.2 | ENST00000234420 | 600678 | Endometrial cancer, familial (AD) Lynch syndrome 5 (AD) Mismatch cancer repair syndrome 3 (AR) | Adult solid tumours cancer susceptibility; Brain cancer pertinent cancer susceptibility; Childhood solid tumours; Haematological malignancies cancer susceptibility; Inherited MMR deficiency (Lynch syndrome); Inherited ovarian cancer (without breast cancer); Inherited polyposis and early onset colorectal cancer - germline testing; Inherited prostate cancer; Ovarian cancer pertinent cancer susceptibility |
| MUTYH | NM_001128425.1 | ENST00000450313 | 604933 | Adenomas, multiple colorectal (AR) | Adult solid tumours cancer susceptibility; Inherited polyposis and early onset colorectal cancer - germline testing |
| NBN | NM_002485.4 | ENST00000265433 | 602667 | Aplastic anemia (NG) Leukemia, acute lymphoblastic (NG) Nijmegen breakage syndrome (AR) | Childhood solid tumours; Haematological malignancies cancer susceptibility; Nijmegen breakage syndrome |
| NF1 | NM_001042492.2 | ENST00000358273 | 613113 | Leukemia, juvenile myelomonocytic (AD) Neurofibromatosis-Noonan syndrome (AD) Neurofibromatosis, familial spinal (AD) Neurofibromatosis, type 1 (AD) Watson syndrome (AD) | Adult solid tumours cancer susceptibility; Childhood solid tumours; Haematological malignancies cancer susceptibility; Neurofibromatosis type 1 (GMS) |
| NF2 | NM_000268.3 | ENST00000338641 | 607379 | Schwannomatosis, vestibular (AD) | Adult solid tumours cancer susceptibility; Childhood solid tumours; Familial tumours of the nervous system |
| NSD1 | NM_022455.4 | ENST00000439151 | 117550 | Sotos syndrome (AD) | Childhood solid tumours |
| NTHL1 | NM_002528.5 | ENST00000219066 | 602656 | Familial adenomatous polyposis 3 (AR) | Adult solid tumours cancer susceptibility; Inherited polyposis and early onset colorectal cancer - germline testing |
| PALB2 | NM_024675.3 | ENST00000261584 | 610355 | Breast-ovarian cancer, familial, susceptibility to, 5 (AD) Pancreatic cancer, susceptibility to, 3 (AD) Fanconi anemia, complementation group N (AR) | Adult solid tumours cancer susceptibility; Breast cancer pertinent cancer susceptibility; Childhood solid tumours; Confirmed Fanconi anaemia or Bloom syndrome; Haematological malignancies cancer susceptibility; Inherited breast cancer and ovarian cancer; Inherited ovarian cancer (without breast cancer); Inherited pancreatic cancer; Inherited prostate cancer |
| PAX5 | NM_016734.2 | ENST00000358127 | 167414 | Leukemia, acute lymphoblastic, susceptibility to, 3 (NG) | Haematological malignancies cancer susceptibility; Inherited susceptibility to acute lymphoblastoid leukaemia (ALL) |
| PDGFRA | NM_006206.4 | ENST00000257290 | 173490 | Gastrointestinal stromal tumor / GIST-plus syndrome, familial (NG) Hypereosinophilic syndrome, idiopathic, resistant to imatinib (IC) | Childhood solid tumours; Inherited predisposition to GIST |
| PHOX2B | NM_003924.3 | ENST00000226382 | 603851 | Neuroblastoma, susceptibility to, 2 (NG) Central hypoventilation syndrome, congenital, 1, with or without Hirschsprung disease (AD) Neuroblastoma with Hirschsprung disease (NG) | Childhood solid tumours |
| PMS2 | NM_000535.5 | ENST00000265849 | 600259 | Lynch syndrome 4 (NG) Mismatch cancer repair syndrome 4 (AR) | Adult solid tumours cancer susceptibility; Brain cancer pertinent cancer susceptibility; Childhood solid tumours; Haematological malignancies cancer susceptibility; Inherited MMR deficiency (Lynch syndrome); Inherited polyposis and early onset colorectal cancer - germline testing |
| POLD1 | NM_002691.3 | ENST00000440232 | 174761 | Colorectal cancer, susceptibility to, 10 (AD) Immunodeficiency 120 (AR) Mandibular hypoplasia, deafness, progeroid features, and lipodystrophy syndrome (AD) | Adult solid tumours cancer susceptibility; APC associated Polyposis; Brain cancer pertinent cancer susceptibility; Childhood solid tumours; Inherited polyposis and early onset colorectal cancer - germline testing |
| POLE | NM_006231.2 | ENST00000320574 | 174762 | Colorectal cancer, susceptibility to, 11 (AD) FILS syndrome (AR) IMAGE-I syndrome (AR) | Adult solid tumours cancer susceptibility; Inherited polyposis and early onset colorectal cancer - germline testing |
| POT1 | NM_015450.2 | ENST00000357628 | 606478 | ?Cerebroretinal microangiopathy with calcification and cysts 3 (AR) ?Pulmonary fibrosis and/or bone marrow failure syndrome, telomere-related, 8 (AD) Tumor predisposition syndrome 3 (AD) | Familial melanoma; Li Fraumeni Syndrome |
| PPM1D | NM_003620.3 | ENST00000305921 | 605100 | Jansen-de Vries syndrome (AD) |  |
| PRF1 | NM_001083116.1 | ENST00000441259 | 170280 | Aplastic anemia (NG) Hemophagocytic lymphohistiocytosis, familial, 2 (AR) Lymphoma, non-Hodgkin (NG) | Haematological malignancies cancer susceptibility |
| PRKAR1A | NM_002734.3 | ENST00000392710 | 188830 | Acrodysostosis 1, with or without hormone resistance (AD) Carney complex, type 1 (AD) Myxoma, intracardiac (AD) Pigmented nodular adrenocortical disease, primary, 1 (AD) | Carney complex; Childhood solid tumours; Endocrine neoplasia |
| PTCH1 | NM_000264.3 | ENST00000331920 | 601309 | Basal cell nevus syndrome 1 (AD) Holoprosencephaly 7 (AD) | Adult solid tumours cancer susceptibility; Childhood solid tumours; Nevoid Basal Cell Carcinoma Syndrome or Gorlin syndrome |
| PTEN | NM_000314.4 | ENST00000371953 | 601728 | Glioma susceptibility 2 (AD) Meningioma (AD) Cowden syndrome 1 (AD) Lhermitte-Duclos disease (AD) Macrocephaly / autism syndrome (AD) | Adult solid tumours cancer susceptibility; Childhood solid tumours; Inherited polyposis and early onset colorectal cancer - germline testing; PTEN Hamartoma Tumour Syndrome |
| RAD51C | NM_058216.1 | ENST00000337432 | 602774 | Breast-ovarian cancer, familial, susceptibility to, 3 (NG) Fanconi anemia, complementation group O (AR) | Adult solid tumours cancer susceptibility; Inherited breast cancer and ovarian cancer; Inherited ovarian cancer (without breast cancer); Ovarian cancer pertinent cancer susceptibility |
| RAD51D | NM_002878.3 | ENST00000345365 | 602954 | Breast-ovarian cancer, familial, susceptibility to, 4 (NG) | Adult solid tumours cancer susceptibility; Inherited breast cancer and ovarian cancer; Inherited ovarian cancer (without breast cancer); Ovarian cancer pertinent cancer susceptibility |
| RB1 | NM_000321.2 | ENST00000267163 | 614041 | Retinoblastoma / Retinoblastoma, trilateral (AD) | Adult solid tumours cancer susceptibility; Childhood solid tumours; Retinoblastoma |
| RECQL4 | NM_004260.3 | ENST00000428558 | 603780 | Baller-Gerold syndrome (AR) RAPADILINO syndrome (AR) Rothmund-Thompson syndrome, type 2 (AR) | Childhood solid tumours; Sarcoma susceptibility |
| RET | NM_020975.4 | ENST00000355710 | 164761 | Hirschsprung disease, protection against (AD) Hirschsprung disease, susceptibility to, 1 (AD) Medullary thyroid carcinoma (AD) Multiple endocrine neoplasia IIA (AD) Multiple endocrine neoplasia IIB (AD) Pheochromocytoma (AD) | Adult solid tumours cancer susceptibility; Childhood solid tumours; Endocrine neoplasia; Inherited phaeochromocytoma and paraganglioma excluding NF1; Multiple endocrine neoplasia type 2 |
| RHBDF2 | NM_024599.5 | ENST00000313080 | 614404 | Tylosis with esophageal cancer (AD) |  |
| RNF43 | NM_017763.5 | ENST00000577716 | 612482 | Sessile serrated polyposis cancer syndrome (AD) | Inherited polyposis and early onset colorectal cancer - germline testing |
| RUNX1 | NM_001754.4 | ENST00000437180 | 151385 | Leukemia, acute myeloid (AD) Platelet disorder, familial, with associated myeloid malignancy (AD) | Haematological malignancies cancer susceptibility; Inherited predisposition to acute myeloid leukaemia (AML) |
| SBDS | NM_016038.2 | ENST00000246868 | 607444 | Aplastic anemia, susceptibility to (NG) Schwachman-Diamond syndrome 1 (AR) | Haematological malignancies cancer susceptibility |
| SDHA | NM_004168.2 | ENST00000264932 | 600857 | Cardiomyopathy, dilated, 1GG (AR) Mitochondrial complex II deficiency, nuclear type 1 (AR) Neurodegeneration with ataxia and late-onset optic atrophy (AD) Pheochromocytoma / paraganglioma syndrome 5 (AD) | Adult solid tumours cancer susceptibility; Inherited phaeochromocytoma and paraganglioma excluding NF1; Inherited predisposition to GIST |
| SDHAF2 | NM_017841.2 | ENST00000301761 | 613019 | Pheochromocytoma / paraganglioma syndrome 2 (AD) | Adult solid tumours cancer susceptibility; Inherited phaeochromocytoma and paraganglioma excluding NF1 |
| SDHB | NM_003000.2 | ENST00000375499 | 185470 | Gastrointestinal stromal tumor (AD) Mitochondrial complex II deficiency, nuclear type 4 (AR) Paraganglioma and gastric stromal sarcoma (NG) Pheochromocytoma / paraganglioma syndrome 4 (AD) | Adult solid tumours cancer susceptibility; Inherited phaeochromocytoma and paraganglioma excluding NF1; Inherited predisposition to GIST; Inherited renal cancer |
| SDHC | NM_003001.3 | ENST00000367975 | 602413 | Gastrointestinal stromal tumor (AD) Paraganglioma and gastric stromal sarcoma (NG) Pheochromocytoma / paraganglioma syndrome 3 (AD) | Adult solid tumours cancer susceptibility; Inherited phaeochromocytoma and paraganglioma excluding NF1; Inherited predisposition to GIST |
| SDHD | NM_003002.2 | ENST00000375549 | 602690 | Mitochondrial complex II deficiency, nuclear type 3 (AR) Paraganglioma and gastric stromal sarcoma (NG) Pheochromocytoma / paraganglioma syndrome 1 (AD) | Adult solid tumours cancer susceptibility; Inherited phaeochromocytoma and paraganglioma excluding NF1; Inherited predisposition to GIST |
| SLX4 | NM_032444.2 | ENST00000294008 | 613278 | Fanconi anemia, complementation group P (AR) | Childhood solid tumours; Confirmed Fanconi anaemia or Bloom syndrome; Haematological malignancies cancer susceptibility |
| SMAD4 | NM_005359.5 | ENST00000342988 | 600993 | Juvenile polyposis / hereditary hemorrhagic telangiectasia syndrome (AD) Myhre syndrome (AD) Polyposis, juvenile intestinal (AD) | Adult solid tumours cancer susceptibility; Childhood solid tumours; Inherited polyposis and early onset colorectal cancer - germline testing |
| SMARCA4 | NM_001128849.1 | ENST00000358026 | 603254 | ?Otosclerosis 12 (AD) Rhabdoid tumor predisposition syndrome 2 (AD) Coffin-Siris syndrome 4 (AD) | Adult solid tumours cancer susceptibility; Childhood solid tumours; Familial rhabdoid tumours |
| SMARCB1 | NM_003073.3 | ENST00000263121 | 601607 | Rhabdoid tumor predisposition syndrome 1 (AD) Schwannomatosis-1, susceptibility to (AD) Coffin-Siris syndrome 3 (AD) | Adult solid tumours cancer susceptibility; Childhood solid tumours; Familial rhabdoid tumours; Familial tumours of the nervous system |
| SMARCE1 | NM_003079.4 | ENST00000348513 | 603111 | Meningioma, familial, susceptibility to (AD) Coffin-Siris syndrome 5 (AD) | Familial tumours of the nervous system |
| SPRED1 | NM_152594.3 | ENST00000299084 | 611431 | Legius syndrome (AD) | Neurofibromatosis type 1 (GMS) |
| STK11 | NM_000455.4 | ENST00000326873 | 602216 | Peutz-Jeghers syndrome (AD) | Adult solid tumours cancer susceptibility; Childhood solid tumours; Inherited pancreatic cancer; Inherited polyposis and early onset colorectal cancer - germline testing |
| SUFU | NM_016169.3 | ENST00000369902 | 607035 | Medulloblastoma (AD, AR) Meningioma, familial, susceptibility to (AD) Basal cell nevus syndrome 2 (NG) Joubert syndrome 32 (AR) | Adult solid tumours cancer susceptibility; Childhood solid tumours; Familial tumours of the nervous system; Nevoid Basal Cell Carcinoma Syndrome or Gorlin syndrome |
| TERT | NM_198253.2 | ENST00000310581 | 187270 | Leukemia, acute myeloid (AD) Melanoma, cutaneous malignant, 9 (AD) Dyskeratosis congenita, autosomal dominant 2 (AD) Dyskeratosis congenita, autosomal recessive 4 (AR) Pulmonary fibrosis and / or bone marrow failure, telomere-related, 1 (AD) | Adult solid tumours cancer susceptibility; Childhood solid tumours; Haematological malignancies cancer susceptibility; Inherited predisposition to acute myeloid leukaemia (AML) |
| TMEM127 | NM_017849.3 | ENST00000258439 | 613403 | Pheochromocytoma, susceptibility to (AD) | Adult solid tumours cancer susceptibility; Inherited phaeochromocytoma and paraganglioma excluding NF1 |
| TP53 | NM_000546.4 | ENST00000269305 | 191170 | Adrenocortical carcinoma, pediatric (AD) Basal cell carcinoma 7 (AD) Choroid plexus papilloma (AD) Colorectal cancer (AD) Glioma susceptibility 1 (AD) Bone marrow failure syndrome 5 (AD) Li-Fraumeni syndrome (AD) | Adult solid tumours cancer susceptibility; Brain cancer pertinent cancer susceptibility; Breast cancer pertinent cancer susceptibility; Childhood solid tumours; Haematological malignancies cancer susceptibility; Inherited predisposition to acute myeloid leukaemia (AML); Li Fraumeni Syndrome; Sarcoma susceptibility |
| TSC1 | NM_000368.4 | ENST00000298552 | 605284 | Lymphangioleiomyomatosis (NG) Tuberous sclerosis-1 (AD) | Adult solid tumours cancer susceptibility; Childhood solid tumours; Tuberous sclerosis |
| TSC2 | NM_000548.3 | ENST00000219476 | 613254 | Tuberous sclerosis-2 (AD) | Adult solid tumours cancer susceptibility; Childhood solid tumours; Tuberous sclerosis |
| VHL | NM_000551.2 | ENST00000256474 | 608527 | Erythrocytosis, familial, 2 (AR) Pheochromocytoma (AD) von Hippel-Lindau syndrome (AD) | Adult solid tumours cancer susceptibility; Childhood solid tumours; Endocrine neoplasia; Inherited phaeochromocytoma and paraganglioma excluding NF1; Inherited renal cancer; Von Hippel Lindau syndrome |
| WRN | NM_000553.4 | ENST00000298139 | 604611 | Werner syndrome (AR) | Childhood solid tumours |
| WT1 | NM_024426.3 | ENST00000332351 | 607102 | Denys-Drash syndrome (AD) Frasier syndrome (AD) Meacham syndrome (AD) Nephrotic syndrome, type 4 (AD) Wilms tumor, type 1 (AD) | Adult solid tumours cancer susceptibility; Childhood solid tumours; Wilms tumour with features suggestive of predisposition |
| XPA | NM_000380.3 | ENST00000375128 | 611153 | Xeroderma pigmentosum, group A (AR) | Adult solid tumours cancer susceptibility; Childhood solid tumours |
| XPC | NM_004628.4 | ENST00000285021 | 613208 | Xeroderma pigmentosum, group C (AR) | Adult solid tumours cancer susceptibility; Childhood solid tumours |

**Table S2.** *Functional, splicing and genetic epidemiological datasets collated in CanVar-UK.* Displayed are all (a) functional studies, including multiplex assays of variant effect (MAVEs), (b) splicing assays and (c) multifactorial genetic epidemiology studies for which variant-level scores are currently housed in CanVar-UK. This list was retrieved on 15^th^ March 2026, but is liable to change prior to publication. Note that some functional analyses cover multiple genes, and so may appear multiple times in the table.

**(a) Functional datasets**

| **Authors** | **Publication year** | **Publication title** | **PubMed ID** | **# CanVar variants** |
| --- | --- | --- | --- | --- |
| **ATM** | | | | |
| Kim et al. | 2025 | Functional assessment of all ATM SNVs using prime editing and deep learning | 40580951 | 29,352 |
| **BAP1** | | | | |
| Waters et al. | 2024 | Saturation genome editing of BAP1 functionally classifies somatic and germline variants | 38969833 | 7557 |
| **BRCA1** | | | | |
| Bouwman et al. | 2020 | Functional Categorization of BRCA1 Variants of Uncertain Clinical Significance in Homologous Recombination Repair Complementation Assays | 32546644 | 288 |
| Fernandes et al. | 2019 | Impact of amino acid substitutions at secondary structures in the BRCT domains of the tumor suppressor BRCA1: Implications for clinical annotation | 30765603 | 398 |
| Petitalot et al. | 2019 | Combining Homologous Recombination and Phosphopeptide-binding Data to Predict the Impact of BRCA1 BRCT Variants on Cancer Risk | 30257991 | 90 |
| Findlay et al. | 2018 | Accurate classification of BRCA1 variants with saturation genome editing | 30209399 | 3893 |
| Starita et al. | 2018 | A Multiplex Homology-Directed DNA Repair Assay Reveals the Impact of More Than 1,000 BRCA1 Missense Substitution Variants on Protein Function | 30219179 | 1093 |
| Bouwman et al. | 2013 | A high-throughput functional complementation assay for classification of BRCA1 missense variants | 23867111 | 86 |
| **BRCA2** | | | | |
| Huang et al. | 2025 | Functional evaluation and classification of BRCA2 variants | 39779857 | 6959 |
| Sahu et al. | 2025 | Saturation genome editing-based clinical classification of BRCA2 variants | 39779848 | 6551 |
| Hu et al. | 2024 | Functional analysis and clinical classification of 462 germline BRCA2 missense variants affecting the DNA binding domain | 38417439 | 510 |
| Hu et al. | 2022 | Classification of BRCA2 Variants of Uncertain Significance (VUS) Using an ACMG/AMP Model Incorporating a Homology-Directed Repair (HDR) Functional Assay | 35736817 | 144 |
| Richardson et al. | 2021 | Strong functional data for pathogenicity or neutrality classify BRCA2 DNA-binding-domain variants of uncertain significance | 33609447 | 275 |
| Biswas et al. | 2020 | A computational model for classification of BRCA2 variants using mouse embryonic stem cell-based functional assays | 33293522 | 101 |
| Ikegami et al. | 2020 | High-throughput functional evaluation of BRCA2 variants of unknown significance | 32444794 | 283 |
| Hart et al. | 2019 | Comprehensive annotation of BRCA1 and BRCA2 missense variants by functionally validated sequence-based computational prediction models | 29884841 | 229 |
| Mesman et al. | 2019 | The functional impact of variants of uncertain significance in BRCA2 | 29988080 | 43 |
| Guidugli et al. | 2018 | Assessment of the Clinical Relevance of BRCA2 Missense Variants by Functional and Computational Approaches | 29394989 | 139 |
| **MLH1** | | | | |
| Bouvet et al. | 2019 | Methylation Tolerance-Based Functional Assay to Assess Variants of Unknown Significance in the MLH1 and MSH2 Genes and Identify Patients With Lynch Syndrome | 30998989 | 44 |
| Drost et al. | 2019 | A functional assay-based procedure to classify mismatch repair gene variants in Lynch syndrome | 30504929 | 38 |
| Drost et al. | 2010 | A cell-free assay for the functional analysis of variants of the mismatch repair protein MLH1 | 20020535 | 26 |
| **MSH2** | | | | |
| Jia et al. | 2021 | Massively parallel functional testing of MSH2 missense variants conferring Lynch syndrome risk | 33357406 | 5872 |
| Bouvet et al. | 2019 | Methylation Tolerance-Based Functional Assay to Assess Variants of Unknown Significance in the MLH1 and MSH2 Genes and Identify Patients With Lynch Syndrome | 30998989 | 44 |
| Drost et al. | 2019 | A functional assay-based procedure to classify mismatch repair gene variants in Lynch syndrome | 30504929 | 35 |
| Drost et al. | 2012 | A rapid and cell-free assay to test the activity of lynch syndrome-associated MSH2 and MSH6 missense variants | 22102614 | 20 |
| **MSH6** | | | | |
| Drost et al. | 2012 | A rapid and cell-free assay to test the activity of lynch syndrome-associated MSH2 and MSH6 missense variants | 22102614 | 20 |
| **MUTYH** | | | | |
| Hemker et al. | 2025 | Saturation mapping of MUTYH variant effects using DNA repair reporters | 40738107 | 4308 |
| **PALB2** | | | | |
| Boonen et al. | 2026 | Site-saturation functional screens identify PALB2 missense variants associated with increased breast cancer risk | 41554690 | 2539 |
| **PTEN** | | | | |
| Mighell, Evans-Dutson & O'Roak | 2018 | A Saturation Mutagenesis Approach to Understanding PTEN Lipid Phosphatase Activity and Genotype-Phenotype Relationships | 29706350 | 2786 |
| **RAD51C** | | | | |
| Olvera-León et al. | 2024 | High-resolution functional mapping of RAD51C by saturation genome editing | 39299233 | 3849 |
| **TP53** | | | | |
| Giacomelli et al. | 2018 | Mutational processes shape the landscape of TP53 mutations in human cancer | 30224644 | 8258 |
| Kato et al. | 2003 | Understanding the function-structure and function-mutation relationships of p53 tumor suppressor protein by high-resolution missense mutation analysis | 12826609 | 2320 |
| **VHL** | | | | |
| Buckley et al. | 2024 | Saturation genome editing maps the functional spectrum of pathogenic VHL alleles | 38969834 | 2268 |

**(b) Splicing datasets**

| **Authors** | **Publication year** | **Publication title** | **PubMed ID** | **# CanVar variants** |
| --- | --- | --- | --- | --- |
| **BRCA1** | | | | |
| Walker et al. | 2013 | Evaluation of a 5-tier scheme proposed for classification of sequence variants using bioinformatic and splicing assay data: inter-reviewer variability and promotion of minimum reporting guidelines | 23893897 | 252 |
| Houdayer et al. | 2012 | Guidelines for splicing analysis in molecular diagnosis derived from a set of 327 combined in silico/in vitro studies on BRCA1 and BRCA2 variants | 22505045 | 113 |
| **BRCA2** | | | | |
| Walker et al. | 2013 | Evaluation of a 5-tier scheme proposed for classification of sequence variants using bioinformatic and splicing assay data: inter-reviewer variability and promotion of minimum reporting guidelines | 23893897 | 194 |
| Houdayer et al. | 2012 | Guidelines for splicing analysis in molecular diagnosis derived from a set of 327 combined in silico/in vitro studies on BRCA1 and BRCA2 variants | 22505045 | 159 |
| **Multiple genes** | | | | |
| Wai et al. | 2020 | Blood RNA analysis can increase clinical diagnostic rate and resolve variants of uncertain significance | 32123317 | 99 |
| Wessex Clinical Genetics Service | Data obtained from NHS clinical testing | | | 112 |

**(c) Multifactorial genetic epidemiological studies**

| **Authors** | **Publication year** | **Publication title** | **PubMed ID** | **# CanVar variants** |
| --- | --- | --- | --- | --- |
| **BRCA1** | | | | |
| Li et al. | 2020 | Classification of variants of uncertain significance in BRCA1 and BRCA2 using personal and family history of cancer from individuals in a large hereditary cancer multigene panel testing cohort | 31853058 | 1302 |
| Parsons et al. | 2019 | Large scale multifactorial likelihood quantitative analysis of BRCA1 and BRCA2 variants: An ENIGMA resource to support clinical variant classification | 31131967 | 523 |
| Vallée et al. | 2019 | Classification of missense substitutions in the BRCA genes: a database dedicated to Ex-UVs | 21990165 | 113 |
| Easton et al. | 2007 | A systematic genetic assessment of 1,433 sequence variants of unknown clinical significance in the BRCA1 and BRCA2 breast cancer-predisposition genes | 17924331 | 105 |
| **BRCA2** | | | | |
| Li et al. | 2020 | Classification of variants of uncertain significance in BRCA1 and BRCA2 using personal and family history of cancer from individuals in a large hereditary cancer multigene panel testing cohort | 31853058 | 2167 |
| Parsons et al. | 2019 | Large scale multifactorial likelihood quantitative analysis of BRCA1 and BRCA2 variants: An ENIGMA resource to support clinical variant classification | 31131967 | 872 |
| Vallée et al. | 2019 | Classification of missense substitutions in the BRCA genes: a database dedicated to Ex-UVs | 21990165 | 104 |
| Easton et al. | 2007 | A systematic genetic assessment of 1,433 sequence variants of unknown clinical significance in the BRCA1 and BRCA2 breast cancer-predisposition genes | 17924331 | 88 |

**Table S3.** *Complete list of CanVIG-UK survey responses for questions related to CanVar-UK.* Responses to the CanVIG-UK survey are presented both unfiltered and stratified into respondents working as clinical scientists (with a self-identified role of “Qualified clinical scientist” or “Clinical scientist trainee”). The survey was available online for all CanVIG-UK members from 14^th^-28^th^ November 2025.

| **Question** | **Option** | **Ranking/score** | **Number (%) of respondents** | |
| --- | --- | --- | --- | --- |
|  |  |  | **Clinical scientists (n=98)** | **All respondents (n=137)** |
| **“Rank the following resources in order of how useful you find them for assembling evidence for variant interpretation in cancer susceptibility genes.”** | Alamut | 1 | 24 (24.49%) | 26 (18.98%) |
|  |  | 2 | 24 (24.49%) | 26 (18.98%) |
|  |  | 3 | 39 (39.8%) | 59 (43.07%) |
|  |  | 4 | 9 (9.18%) | 18 (13.14%) |
|  |  | 5 | 2 (2.04%) | 8 (5.84%) |
|  | CanVar-UK | 1 | 65 (66.33%) | 91 (66.42%) |
|  |  | 2 | 19 (19.39%) | 29 (21.17%) |
|  |  | 3 | 10 (10.2%) | 12 (8.76%) |
|  |  | 4 | 3 (3.06%) | 4 (2.92%) |
|  |  | 5 | 1 (1.02%) | 1 (0.73%) |
|  | ClinVar | 1 | 5 (5.1%) | 15 (10.95%) |
|  |  | 2 | 49 (50%) | 75 (54.74%) |
|  |  | 3 | 37 (37.76%) | 39 (28.47%) |
|  |  | 4 | 7 (7.14%) | 8 (5.84%) |
|  |  | 5 | 0 (0%) | 0 (0%) |
|  | Franklin | 1 | 4 (4.08%) | 5 (3.65%) |
|  |  | 2 | 3 (3.06%) | 4 (2.92%) |
|  |  | 3 | 6 (6.12%) | 9 (6.57%) |
|  |  | 4 | 43 (43.88%) | 61 (44.53%) |
|  |  | 5 | 42 (42.86%) | 58 (42.34%) |
|  | VarSome | 1 | 0 (0%) | 0 (0%) |
|  |  | 2 | 3 (3.06%) | 3 (2.19%) |
|  |  | 3 | 6 (6.12%) | 18 (13.14%) |
|  |  | 4 | 36 (36.73%) | 46 (33.58%) |
|  |  | 5 | 53 (54.08%) | 70 (51.09%) |
| **“How often on average do you use CanVar-UK for the following actions?”** | Accessing information for variant classification | Never | 0 (0%) | 1 (0.73%) |
|  |  | Daily | 43 (43.88%) | 46 (33.58%) |
|  |  | Weekly | 32 (32.65%) | 39 (28.47%) |
|  |  | Monthly | 9 (9.18%) | 21 (15.33%) |
|  |  | 4-6 times per year | 9 (9.18%) | 15 (10.95%) |
|  |  | 1-3 times per year | 5 (5.1%) | 15 (10.95%) |
|  | Accessing links to external websites (e.g. ClinVar, gnomAD) | Never | 12 (12.24%) | 19 (13.87%) |
|  |  | Daily | 30 (30.61%) | 33 (24.09%) |
|  |  | Weekly | 25 (25.51%) | 34 (24.82%) |
|  |  | Monthly | 14 (14.29%) | 21 (15.33%) |
|  |  | 4-6 times per year | 8 (8.16%) | 14 (10.22%) |
|  |  | 1-3 times per year | 9 (9.18%) | 16 (11.68%) |
|  | Posting questions via the discussion forum | Never | 38 (38.78%) | 61 (44.53%) |
|  |  | Daily | 0 (0%) | 0 (0%) |
|  |  | Weekly | 0 (0%) | 2 (1.46%) |
|  |  | Monthly | 17 (17.35%) | 18 (13.14%) |
|  |  | 4-6 times per year | 14 (14.29%) | 15 (10.95%) |
|  |  | 1-3 times per year | 29 (29.59%) | 40 (29.2%) |
|  | Responding to questions via the discussion forum | Never | 43 (43.88%) | 68 (49.64%) |
|  |  | Daily | 0 (0%) | 0 (0%) |
|  |  | Weekly | 0 (0%) | 2 (1.46%) |
|  |  | Monthly | 15 (15.31%) | 15 (10.95%) |
|  |  | 4-6 times per year | 12 (12.24%) | 14 (10.22%) |
|  |  | 1-3 times per year | 28 (28.57%) | 37 (27.01%) |
|  | Posting proposed variant classifications via the discussion forum | Never | 48 (48.98%) | 74 (54.01%) |
|  |  | Daily | 0 (0%) | 0 (0%) |
|  |  | Weekly | 0 (0%) | 1 (0.73%) |
|  |  | Monthly | 6 (6.12%) | 6 (4.38%) |
|  |  | 4-6 times per year | 14 (14.29%) | 15 (10.95%) |
|  |  | 1-3 times per year | 30 (30.61%) | 40 (29.2%) |
| **“How useful do you find the following CanVar-UK resources? (scale 1-5; 1=Not at all useful, 5=Extremely useful)”** | Discussion forum | 1 (Not at all useful) | 3 (3.06%) | 4 (2.92%) |
|  |  | 2 | 9 (9.18%) | 13 (9.49%) |
|  |  | 3 | 17 (17.35%) | 26 (18.98%) |
|  |  | 4 | 21 (21.43%) | 32 (23.36%) |
|  |  | 5 (Extremely useful) | 48 (48.98%) | 62 (45.26%) |
|  | Variant summary information (first tab) | 1 (Not at all useful) | 1 (1.02%) | 2 (1.46%) |
|  |  | 2 | 7 (7.14%) | 9 (6.57%) |
|  |  | 3 | 12 (12.24%) | 17 (12.41%) |
|  |  | 4 | 40 (40.82%) | 51 (37.23%) |
|  |  | 5 (Extremely useful) | 38 (38.78%) | 58 (42.34%) |
|  | Links to other websites (e.g. ClinVar, gnomAD) | 1 (Not at all useful) | 2 (2.04%) | 2 (1.46%) |
|  |  | 2 | 12 (12.24%) | 16 (11.68%) |
|  |  | 3 | 17 (17.35%) | 25 (18.25%) |
|  |  | 4 | 28 (28.57%) | 42 (30.66%) |
|  |  | 5 (Extremely useful) | 39 (39.8%) | 52 (37.96%) |
|  | PM5 function (REVEL score and ClinVar status for other variants at the same codon) | 1 (Not at all useful) | 1 (1.02%) | 1 (0.73%) |
|  |  | 2 | 2 (2.04%) | 8 (5.84%) |
|  |  | 3 | 14 (14.29%) | 23 (16.79%) |
|  |  | 4 | 29 (29.59%) | 39 (28.47%) |
|  |  | 5 (Extremely useful) | 52 (53.06%) | 66 (48.18%) |
|  | In silico predictions | 1 (Not at all useful) | 2 (2.04%) | 3 (2.19%) |
|  |  | 2 | 1 (1.02%) | 6 (4.38%) |
|  |  | 3 | 15 (15.31%) | 21 (15.33%) |
|  |  | 4 | 29 (29.59%) | 42 (30.66%) |
|  |  | 5 (Extremely useful) | 51 (52.04%) | 65 (47.45%) |
|  | Functional/splicing assay data | 1 (Not at all useful) | 1 (1.02%) | 2 (1.46%) |
|  |  | 2 | 1 (1.02%) | 6 (4.38%) |
|  |  | 3 | 7 (7.14%) | 13 (9.49%) |
|  |  | 4 | 19 (19.39%) | 31 (22.63%) |
|  |  | 5 (Extremely useful) | 70 (71.43%) | 85 (62.04%) |
|  | Genetic epidemiology data | 1 (Not at all useful) | 1 (1.02%) | 1 (0.73%) |
|  |  | 2 | 1 (1.02%) | 4 (2.92%) |
|  |  | 3 | 12 (12.24%) | 18 (13.14%) |
|  |  | 4 | 18 (18.37%) | 30 (21.9%) |
|  |  | 5 (Extremely useful) | 66 (67.35%) | 84 (61.31%) |
|  | UK laboratory case data | 1 (Not at all useful) | 0 (0%) | 1 (0.73%) |
|  |  | 2 | 4 (4.08%) | 7 (5.11%) |
|  |  | 3 | 6 (6.12%) | 14 (10.22%) |
|  |  | 4 | 17 (17.35%) | 26 (18.98%) |
|  |  | 5 (Extremely useful) | 71 (72.45%) | 89 (64.96%) |
|  | Population control data | 1 (Not at all useful) | 2 (2.04%) | 2 (1.46%) |
|  |  | 2 | 2 (2.04%) | 4 (2.92%) |
|  |  | 3 | 15 (15.31%) | 22 (16.06%) |
|  |  | 4 | 28 (28.57%) | 39 (28.47%) |
|  |  | 5 (Extremely useful) | 51 (52.04%) | 70 (51.09%) |
|  | Gene-level information (bubble) | 1 (Not at all useful) | 4 (4.08%) | 5 (3.65%) |
|  |  | 2 | 12 (12.24%) | 16 (11.68%) |
|  |  | 3 | 32 (32.65%) | 42 (30.66%) |
|  |  | 4 | 29 (29.59%) | 40 (29.2%) |
|  |  | 5 (Extremely useful) | 21 (21.43%) | 34 (24.82%) |
|  | Intron/exon information | 1 (Not at all useful) | 3 (3.06%) | 4 (2.92%) |
|  |  | 2 | 16 (16.33%) | 22 (16.06%) |
|  |  | 3 | 27 (27.55%) | 34 (24.82%) |
|  |  | 4 | 28 (28.57%) | 40 (29.2%) |
|  |  | 5 (Extremely useful) | 24 (24.49%) | 37 (27.01%) |
|  | Variant evidence summary PDF generation | 1 (Not at all useful) | 15 (15.31%) | 16 (11.68%) |
|  |  | 2 | 15 (15.31%) | 17 (12.41%) |
|  |  | 3 | 30 (30.61%) | 38 (27.74%) |
|  |  | 4 | 17 (17.35%) | 30 (21.9%) |
|  |  | 5 (Extremely useful) | 21 (21.43%) | 36 (26.28%) |
| **“Overall, how important is CanVar-UK for your work?**  **(scale 0-10; 0=Not at all, 10=Essential)?”** | | 0 | 0 (0%) | 1 (0.73%) |
|  |  | 1 | 0 (0%) | 1 (0.73%) |
|  |  | 2 | 1 (1.02%) | 2 (1.46%) |
|  |  | 3 | 1 (1.02%) | 2 (1.46%) |
|  |  | 4 | 0 (0%) | 0 (0%) |
|  |  | 5 | 5 (5.1%) | 9 (6.57%) |
|  |  | 6 | 1 (1.02%) | 6 (4.38%) |
|  |  | 7 | 5 (5.1%) | 8 (5.84%) |
|  |  | 8 | 12 (12.24%) | 19 (13.87%) |
|  |  | 9 | 18 (18.37%) | 24 (17.52%) |
|  |  | 10 | 55 (56.12%) | 65 (47.45%) |
